# Post-acute health care burden after SARS-CoV-2 infection: A retrospective cohort study of long COVID among 530,892 adults

**DOI:** 10.1101/2022.05.06.22274782

**Authors:** Candace D. McNaughton, Peter C. Austin, Atul Sivaswamy, Jiming Fang, Husam Abdel-Qadir, Nick Daneman, Jacob A. Udell, Walter Wodchis, Ivona Mostarac, Clare L. Atzema

## Abstract

**Importance:** The SARS-CoV-2 pandemic portends a significant increase in health care use related to post-acute COVID sequelae, but the magnitude is not known.

**Objective:** To assess the burden of post-acute health care use after a positive versus negative polymerase chain reaction (PCR) test for SARS-CoV-2.

**Design, Setting, and Participants:** Retrospective cohort study of community-dwelling adults January 1, 2020 to March 31, 2021 in Ontario, Canada, using linked population-based health data. Follow-up began 56 days after PCR testing.

**Exposures:** Individuals with a positive SARS-CoV-2 PCR test were matched 1:1 to individuals who tested negative based on hospitalization, test date, public health unit, sex, and a propensity score of socio-demographic and clinical characteristics.

**Main Outcomes and Measures:** The health care utilization rate was the number of outpatient clinical encounters, homecare encounters, emergency department visits, days hospitalized, and days in long-term care per person-year. Mean health care utilization for test-positive versus negative individuals was compared using negative binomial regression, and rates at 95^th^ and 99^th^ percentiles were compared. Outcomes were also stratified by sex.

**Results:** Among 530,232 unique, matched individuals, mean age was 44 years (sd 17), 51% were female, and 0.6% had received ≥1 COVID-19 vaccine dose. The mean rate of health care utilization was 11% higher in test-positive individuals (RR 1.11, 95% confidence interval [CI] 1.10-1.13). At the 95^th^ percentile, test-positive individuals had 2.1 (95% CI 1.5-2.6) more health care encounters per person-year, and at the 99^th^ percentile 71.9 (95% CI 57.6-83.2) more health care encounters per person-year. At the 95^th^ percentile, test-positive women had 3.8 (95% CI 2.8-4.8) more health care encounters per person-year while there was no difference for men. At the 99^th^ percentile, test-positive women had 76.7 (95% CI 56.3-89.6) more encounters per person-year, compared to 37.6 (95% CI 16.7-64.3) per person-year for men.

**Conclusions and Relevance:** Post-acute health care utilization after a positive SARS-CoV-2 PCR test is significantly higher compared to matched test-negative individuals. Given the number of infections worldwide, this translates to a tremendous increase in use of health care resources. Stakeholders can use these findings to prepare for health care demand associated with long COVID.

**Key Points:** *Question:* How does the burden of health care use ≥56 days after a positive SARS-CoV-2 polymerase chain reaction (PCR) test compare to matched individuals who tested negative?

*Findings:* After accounting for multiple factors, the mean burden of post-acute health care use was 11% higher among those who tested positive, with higher rates of outpatient encounters, days hospitalized, and days in long-term care. Rates of homecare use were higher for test-positive women but lower for men. For perspective, for every day in January 2022 with 100,000 or more infections, this translates to an estimated 72,000 *additional* post-acute health care encounters per year for the 1% of people who experienced the most severe complications of SARS-CoV-2; among those in the top 50% of health care use, this translates to 245,000 *additional* health care encounters per year. This increase will occur in the context of an ongoing pandemic and, in many health care systems, a depleted workforce and backlogs of care. Unless addressed, this increase is likely to exacerbate existing health inequities.

*Meaning:* Given the large number of people infected, stakeholders can use these findings to plan for health care use associated with long COVID.

## Introduction/Background

The public health implications of the coronavirus 2019 (COVID-19) pandemic caused by airborne spread of the severe acute respiratory syndrome coronavirus 2 (SARS-CoV-2) are difficult to overstate.^1^ There have been >400 million SARS-CoV-2 infections and 5.9 million deaths reported worldwide,^2^ likely gross underestimates as only 3-35% of infections are detected.^3^

In addition to acute illness, there is accumulating evidence that SARS-CoV-2 can cause long-term morbidity.^4-9^ Of those discharged from the hospital after COVID-19, up to 27% die or are re-hospitalized within 60 days, and as many as 70% of non-hospitalized patients report at least one symptom four months after initial infection.^10,11^ Disease severity assessed by mortality and acute hospitalizations alone underestimates the burden of disease caused by SARS-CoV-2 infection.^12,13^

Health care funders, policy makers, and clinicians need a clear understanding of post-acute health care use following SARS-CoV-2 infection, i.e., long COVID, in order to equitably allocate resources now and plan for future needs.^14^ Therefore, we sought to quantify, describe, and compare the post-acute burden of health care use associated with SARS-CoV-2 to negative controls among community-dwelling adults for the entire province of Ontario, Canada.

## Methods

### Study Design

This retrospective cohort study was performed at ICES, previously the Institute for Clinical Evaluative Sciences, which is an independent, non-profit research institute funded by the Ontario Ministry of Health and Long-Term Care and has legal status under Ontario’s Personal Health Information Protection Act (section 45) allowing collection and analysis of health care and demographic data, without consent, for health system evaluation and improvement.^15^

### Cohort creation

We constructed a retrospective cohort of all adults who underwent PCR testing for SARS-CoV-2 between January 1, 2020 and March 31, 2021 in Ontario, Canada. A full list of datasets is in Table E1. All testing was performed within the health care system of Ontario, which is administered by the Ontario Health Insurance Plan (OHIP) and provides publicly funded physician and hospital services for the 14.8 million residents of Ontario.

The date of the first outpatient PCR test that detected SARS-CoV-2 was used as the index date for exposed individuals. For individuals with multiple negative PCR tests, the index date was last test date. Adults (≥18 years of age) who were alive eight weeks (56 days) after their index date were included. Individuals residing in long-term care facilities on their index date were excluded, as were those without a valid date of birth, sex, or death information.

### Exposure definition

Individuals were categorized according to results of SARS-CoV-2 PCR testing as test-negative or test-positive. Pending or indeterminate test results (<0.02%) were excluded.

### Matching

Individuals with a positive SARS-CoV-2 PCR test result were matched to those with only negative test results by sex, hospitalization within two weeks of the index date, test date, public health unit, and a propensity score computed from myriad factors including health care utilization in the previous year, age, baseline socio-demographics and clinical characteristics, neighborhood level socioeconomic indices, and vaccination status (Table E2).^16^ Subjects were matched on the logit of the propensity score using a calliper width equal to 0.05 times the standard deviation of the propensity score.^17^ A standardized difference <0.1 in baseline characteristics was considered a good match.^16,18^

### Outcome definitions

Follow-up to assess post-acute health care utilization began ≥8 weeks (≥56 days) after the index SARS-CoV-2 PCR test date. As the definition of post-acute COVID-19 syndrome, or long COVID, continues to evolve, this timeframe was chosen based on the duration of typical SARS-CoV-2 infectivity and acute symptoms.^1,19-21^ The primary outcome was health care use rate, a composite measure of health care utilization per person-year of follow-up time. Healthcare utilization was the sum of the counts of outpatient encounters (in-person, phone, and virtual), emergency department visits not resulting in hospitalization, days hospitalized, homecare visits (e.g., wound care), and days in long-term care. Each component of health care utilization was examined separately in pre-planned secondary analyses. Follow-up ended on September 30, 2021 or death, whichever occurred first. Sensitivity analyses required hospital discharge to start follow-up, censored at entry into long-term care, censored at six months, and matched by intensive care unit admission.

### Statistical Analyses

All analyses were conducted in SAS version 9.04 at the patient level. Baseline characteristics were reported as means with standard deviations (SD), medians and interquartile ranges (IQR), or frequencies, as appropriate. Given the skewed distribution of health care utilization, this was reported as means and SDs, as well as median, 95^th^ percentile, and 99^th^ percentile. Outcomes were also stratified by sex (male/female).^22,23^

The mean rates of composite and component health care utilization were compared in the matched sample between test-positive and test-negative individuals using univariate negative binomial regression modelling, accounting for matched pairs.^24^ The 95^th^ and 99^th^ percentiles of the person-specific rate of health care utilization for matched test-positive versus negative individuals were compared^25^ with 95% CI’s generated by bootstrapping with 1000 replicates.^26^ Comparisons were also stratified by sex.

## Results

Between January 1, 2020 and March 31, 2021 there were >11 million SARS-CoV-2 PCR tests completed in 3,777,451 unique adults (Figure E1). Of the 3,631,040 individuals who met all inclusion criteria 268,521 (7.4%) had a positive SARS-CoV-2 PCR test. One-to-one matching was successful for 99% of these, thus the matched cohort consists of 530,232 individuals. Demographics, clinical characteristics, and standardized differences between positive and negative individuals for the matched and unmatched cohorts are reported in Table 1 and Table E3, respectively. Compared to the unmatched cohort, the matched cohort was slightly younger, had fewer women and lower income individuals, was more rural and more ethnically diverse, and a greater proportion underwent PCR testing during late 2020 or early 2021.

**Table 1:**
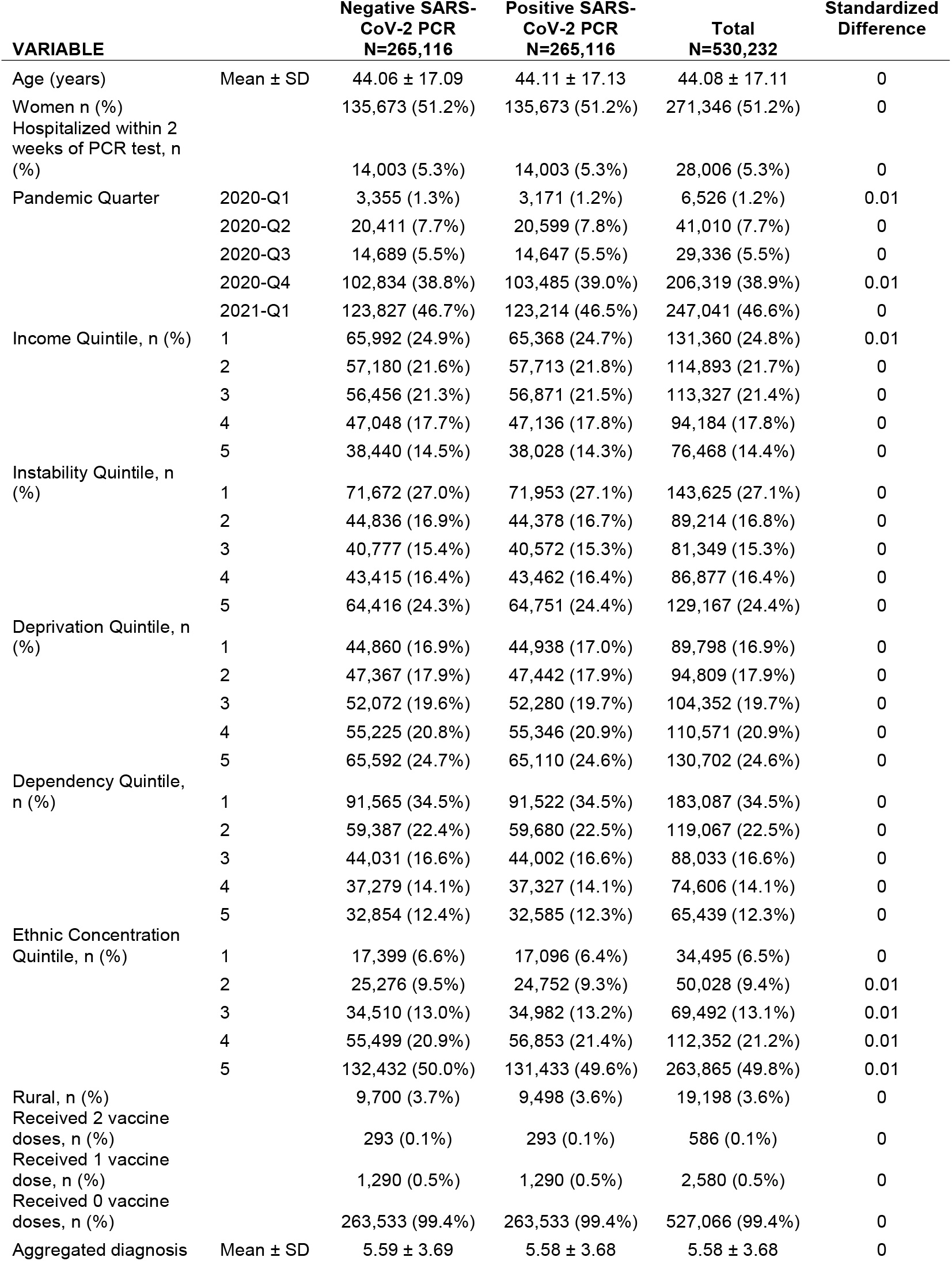
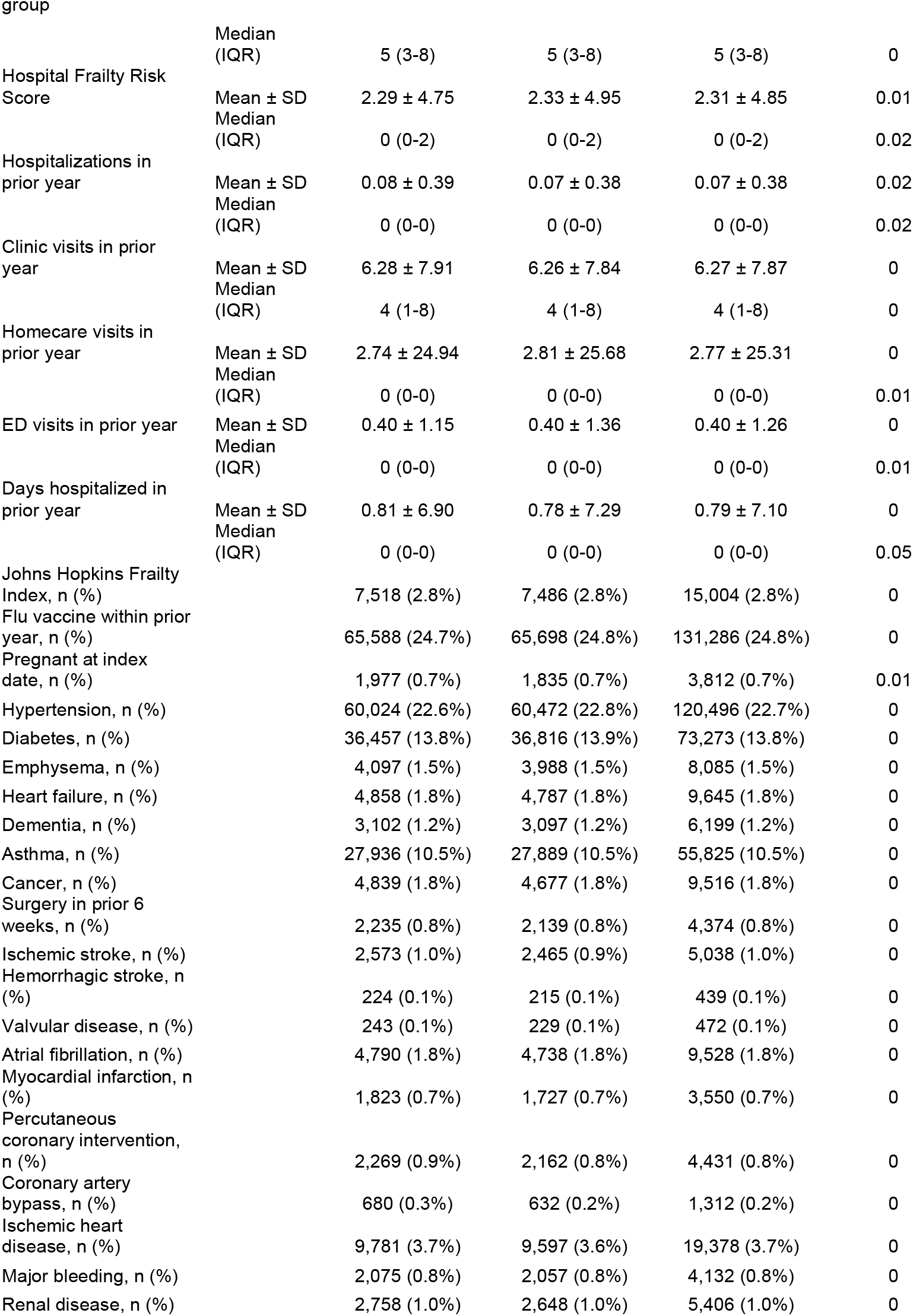
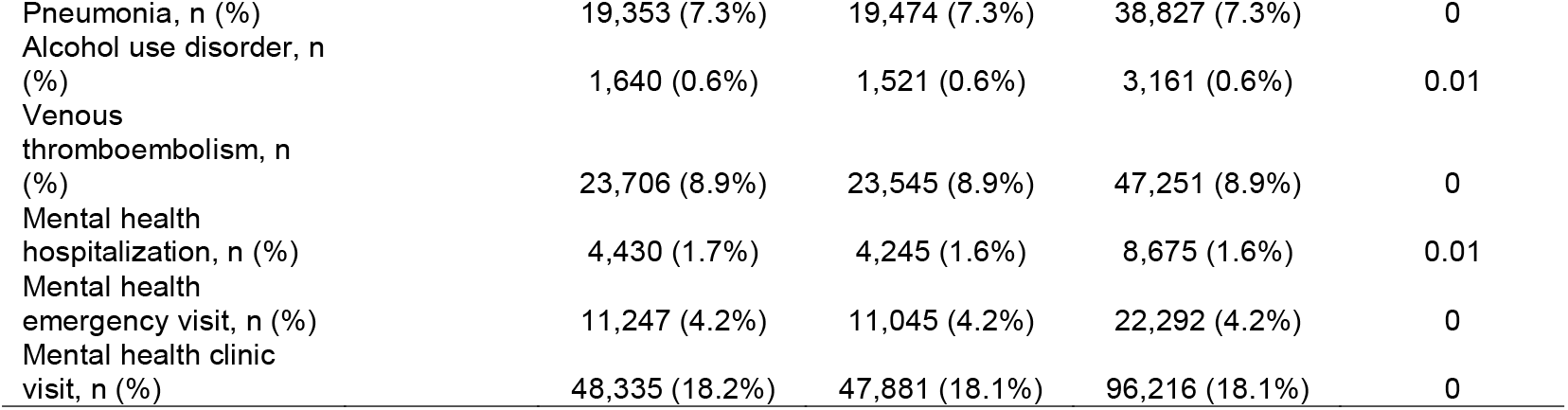
Baseline demographics and clinical characteristics of the matched cohort.

In the matched cohort, mean age was 44 (sd 17) years, 51% were female, and 0.6% had received one or more COVID-19 vaccine doses. Within two weeks after PCR testing, 5.3% were hospitalized in both test-positive and test-negative groups, and pre-existing conditions such as hypertension, diabetes, asthma, prior pneumonia, and prior venous thromboembolism were common. The median number of outpatient clinic encounters prior to the index date was 4 (IQR 1-8) per person-year, with a median of 0 (IQR 0-0) per person-year of homecare visits, emergency department visits, and days hospitalized (Table E4). There was no difference in mortality between groups, including when stratified by sex; mortality was 0.5% for the first six months of follow-up.

### Healthcare utilization

Person-specific health care utilization rates and follow-up, overall and stratified by sex, are shown in Table 2. Median follow-up was 221 days (IQR 187-267) for PCR test-positive and 221 (IQR 188-267) for test-negative individuals. The absolute increase in mean person-specific rate of health care utilization was 1.4 additional encounters (95% CI 1.2-1.6) per person-year for matched test-positive versus test-negative individuals. Using a negative binomial model (Table 3 and Panel A in Figure 1), this was equivalent to a relative increase of 11% for test-positive versus test-negative individuals (RR 1.11, 95% CI 1.09-1.13). Relative increases in types of health care ranged from 3% for emergency department visits and 5% for outpatient clinic encounters, up to 49% for mean days hospitalized (RR 1.49, 95% CI, 1.41-1.57) and 255% for days in long-term care (RR 2.55, 95% CI, 2.28-2.86). In the overall matched cohort, there was no detectible difference in mean homecare encounters by test positivity.

**Table 2:**
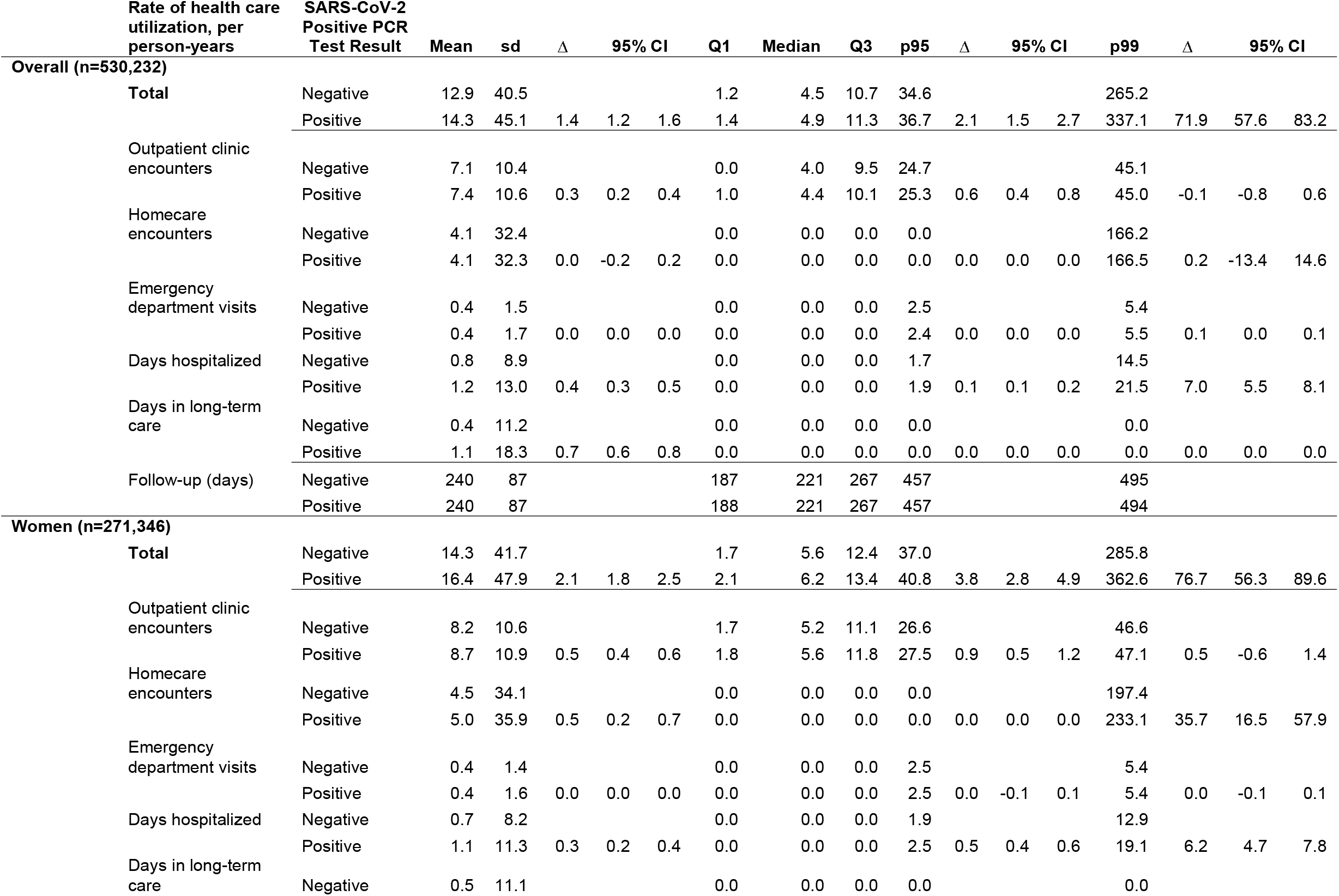
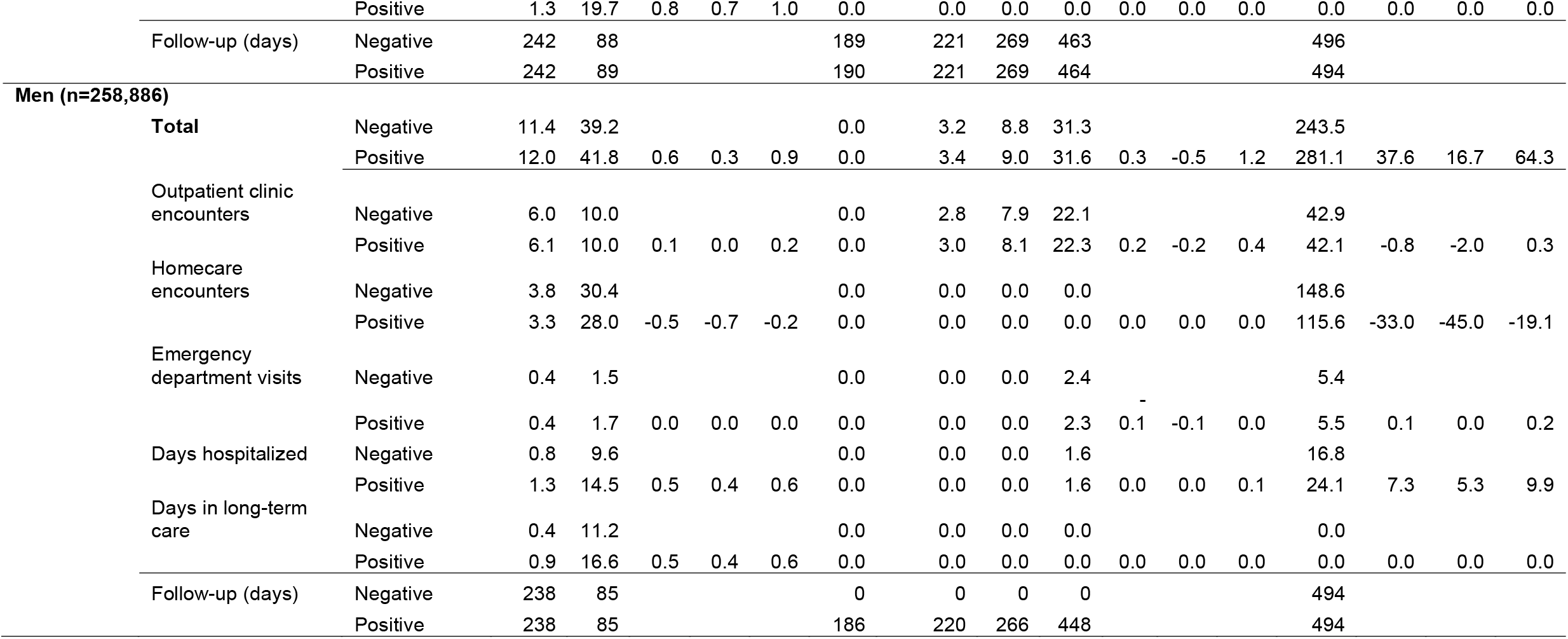
**Total and component health care utilization ≥56 days after SARS-CoV-2 PCR, test-positive versus test-negative individuals, and distribution of follow-up time. Healthcare utilization rates reported per person-year, overall and stratified by sex. The difference in overall health care utilization rates between test-positive and negative individuals are reported at the 95th and 99th percentiles, with their corresponding 95% CI**.

**Table 3:** Rate ratios* of composite and component health care utilization, overall and stratified by sex, for SARS-CoV-2 PCR test-positive versus negative individuals.

| **Healthcare Utilization** | **RR** | **95% CI** |  | **p-value** |
| --- | --- | --- | --- | --- |
| **Overall** |  |  |  |  |
| Total Healthcare Utilization | 1.11 | 1.09 | 1.13 | <.0001 |
| Outpatient clinic encounters | 1.05 | 1.04 | 1.05 | <.0001 |
| Homecare encounters | 1.00 | 0.96 | 1.05 | 0.82 |
| Emergency department visits | 1.03 | 1.02 | 1.05 | 0.0004 |
| Days hospitalized | 1.49 | 1.41 | 1.57 | <.0001 |
| Days in long-term care | 2.55 | 2.28 | 2.86 | <.0001 |
| **Women** |  |  |  |  |
| Total Healthcare Utilization | 1.15 | 1.13 | 1.18 | <.0001 |
| Outpatient clinic encounters | 1.06 | 1.05 | 1.07 | <.0001 |
| Homecare encounters | 1.11 | 1.05 | 1.17 | 0.0003 |
| Emergency department visits | 1.04 | 1.02 | 1.07 | 0.0015 |
| Days hospitalized | 1.42 | 1.32 | 1.52 | <.0001 |
| Days in long-term care | 2.88 | 2.47 | 3.36 | <.0001 |
| **Men** |  |  |  |  |
| Total Healthcare Utilization | 1.06 | 1.03 | 1.08 | <.0001 |
| Outpatient clinic encounters | 1.02 | 1.01 | 1.03 | 0.0003 |
| Homecare encounters | 0.88 | 0.82 | 0.93 | <.0001 |
| Emergency department visits | 1.03 | 1.00 | 1.06 | 0.06 |
| Days hospitalized | 1.56 | 1.44 | 1.69 | <.0001 |
| Days in long-term care | 2.18 | 1.84 | 2.59 | <.0001 |

\* point estimate and confidence intervals computed using negative binomial regression models

**Figure 1:**
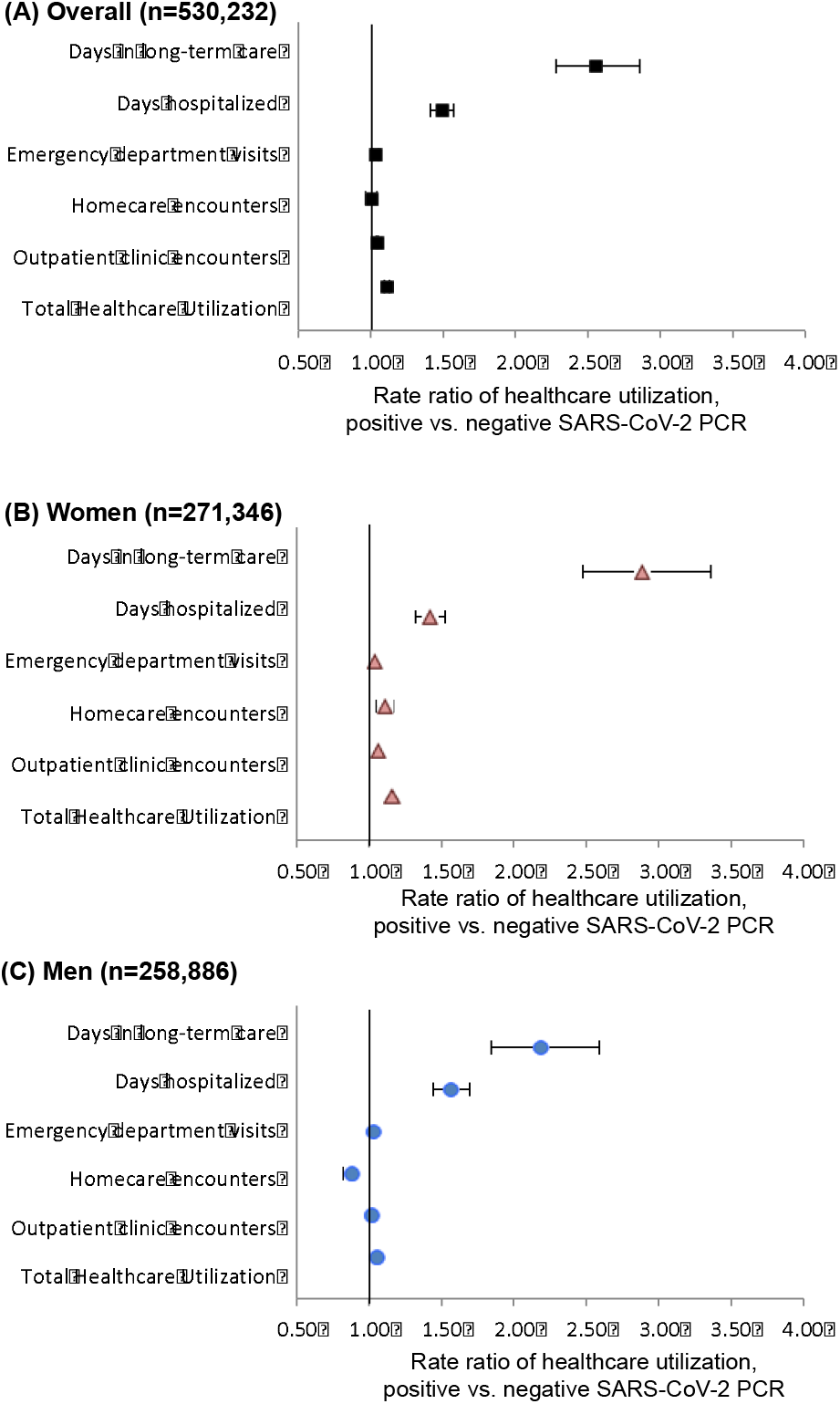
Rate ratios (RR) and 95% confidence intervals for total health care utilization and type of health care encounter, comparing individuals with a positive SARS-CoV-2 PCR test to matched individuals with negative PCR: (A) overall (n=530,232), (B) women (n=271,346), and (C) men (n=258,886).

At the 95^th^ percentile of health care utilization (Table 2, Panel A in Figure E2), test-positive individuals had 2.1 (95% CI 1.5-2.6) additional health care encounters per person-year compared to test-negative individuals. The association of SARS-CoV-2 positivity on the 95^th^ percentile of use was strongest for outpatient encounters, with an absolute increase of 0.6 (95% CI, 0.4-0.8) encounters per person-year compared to matched individuals who tested negative, followed by days hospitalized, with 0.1 (95% CI 0.1-0.2) additional days hospitalized per person-year.

At the 99^th^ percentile of health care utilization (Table 2, Panel A in Figure 2), test-positive individuals had 71.9 (95% CI 57.6-83.2) additional encounters per person-year compared to matched test-negative individuals. The association of SARS-CoV-2 positivity on the 99^th^ percentile of use was strongest for days hospitalized, with an absolute increase of 7.0 (95% CI, 5.5-8.1) days hospitalized per person-year but no differences for outpatient clinic encounters, homecare encounters, emergency department visits, or days in long-term care.

**Figure 2:**
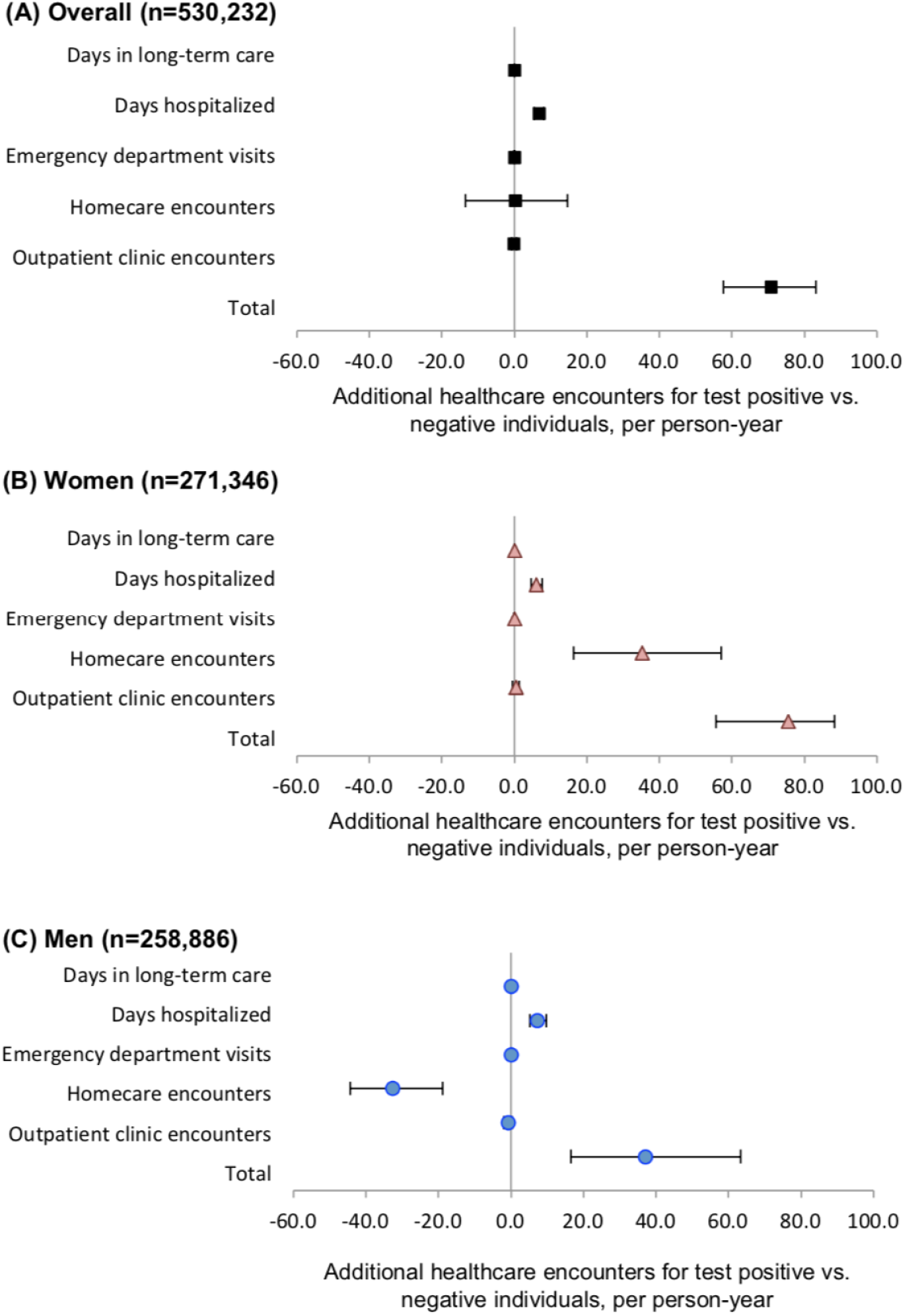
Among individuals at the 99^th^ percentile of health care utilization, additional health care encounters (95% CI) per person-year total and by type of health care encounter, comparing individuals with a positive SARS-CoV-2 PCR test to matched individuals with negative PCR: (A) overall (n=530,232), (B) women (n=271,346), and (C) men (n=258,886).

### Stratified Analyses

Among women (Table 2 and Panel B in Figure 1), on average test-positive women had an absolute increase of 2.1 (95% CI 1.8-2.5) additional encounters per person-year compared to test-negative women, or a RR of 1.15 (95% CI 1.13-1.18; Table 3). At the 95^th^ percentile of health care utilization (Figure E2, Panel B), test-positive women had 3.8 (95% CI 2.8-4.9) additional encounters per person-year, with 0.9 (95% CI, 0.5-1.2) additional outpatient encounters per person-year, 0.5 (95% CI 0.4-0.6) additional days hospitalized per person-year, and no difference in homecare encounters, emergency department visits, or days in long-term care. At the 99^th^ percentile of health care utilization (Figure 2, Panel B), test-positive women had 76.7 (95% CI 56.3-89.6) additional health care encounters per person-year compared to test-negative women, with 35.7 (95% CI 16.5-57.9) additional homecare encounters per person-year, 6.2 (95% CI, 4.7-7.8) additional days hospitalized per person-year, and no detectible difference in outpatient clinic encounters, emergency department visits, or days in long-term care.

In contrast, among men (Table 2 and Panel C in Figure 1), mean health care utilization increased by 0.6 (9% CI 0.3-0.9) encounters per person-year for test-positive versus test-negative men, or a RR of 1.06 (95% CI 1.03-1.08; Table 3). At the 95^th^ percentile of health care utilization (Figure E2, Panel C) there was no detectable difference in composite or components of health care utilization for men. At the 99^th^ percentile of health care utilization (Figure 2, Panel C), test-positive men had 37.6 (95% CI 16.7-64.3) additional health care encounters per person-year, with 7.3 (95% CI 5.3-9.9) additional days hospitalized person-year. Homecare encounters for men decreased by 33.0 (95% CI 19.1-45.0) per person-year, with no difference in outpatient clinic visits, emergency department visits, or days in long-term care.

### Sensitivity Analyses

Overall, findings were robust in sensitivity analyses. Requiring hospital discharge to initiate follow-up (Table E5, Panel A) decreased days hospitalized among individuals at the 99^th^ percentile from 7.0 to 2.2 additional days per person-year. Censoring on entrance to long-term care produced smaller magnitude additional total health care utilization rates and days hospitalized at the mean and 95^th^ percentile, and 10.5 additional homecare encounters per person-year at the 99^th^ percentile health care utilization. Censoring follow-up at six months (Table E5, Panel B) increased magnitude of additional mean health care use (4.4 additional encounters per person-year for test-positive individuals, including mean 6.4 additional days hospitalized per person-year and 4.7 additional days in long-term care per person-year, compared to 1.3, 0.7, and 4.7, per person-year, respectively in main analyses). Results at the 95^th^ percentile were similar to main findings, while the 99^th^ percentile was notable only for fewer homecare encounters. Finally, matching by intensive care unit admission within two weeks after the index date reduced sample size due to fewer successful matches and smaller magnitude additional health care use compared to main analyses.

## Discussion

Our study is unique in its use of population-wide PCR results, outcomes, and socio-demographic data among nearly all adults in the highly diverse Ontario population, where PCR testing was publicly funded, as well as relatively long follow-up time. We found that among 530,232 community-dwelling adults who underwent PCR testing for SARS-CoV-2 between January 2020 and March 2021, individuals with a positive test had 11% higher mean post-acute health care utilization, after accounting for a multitude of factors including acute hospitalization as a measure of acute disease severity, socio-demographic factors, comorbidities, and pandemic wave. This increase was largely driven by a subset of individuals who experienced large increases in health care utilization: test-positive individuals in the 95^th^ percentile of health care use had ∼2 additional encounters per person-year compared to matched controls, while those in the 99^th^ percentile had ∼72 additional health care encounters per person-year. Together with sensitivity analyses, these findings indicate that eight weeks or more after a positive SARS-CoV-2 PCR test, a subset of individuals are not able to live alone at home without support and they utilize substantial health care resources.

These findings portend a tremendous increase in need for health care related to long COVID.^27^ Conservative estimates indicate that 4 million people in Ontario, or 25% of the population, have been recently infected with SARS-CoV-2.^28^ Overall, our findings suggest these will lead to 5.6 million additional health care encounters per year, including 2.9 million additional encounters including 280,000 additional days hospitalized per year for individuals in the highest 1% of health care use. In the United States, where an estimated 140 million people have been recently infected with SARS-CoV-2,^29^ this translates to an additional 196 million additional health care encounters per year, including 10 million health care encounters per year for individuals in the highest 1% of health care use. Increases of this magnitude will require significant restructuring, innovation, and investment of resources, particularly in the context of existing prolonged wait times to access care, insufficient supply of acute and long-term care beds, and projected loss of health care workers.^30-37^

Patterns of health care use differed by sex, adding to known differences by sex related to COVID-19.^38-45^ Among women with a positive SARS-CoV-2 PCR test, increase in total health care utilization was larger than for men (at the 95% percentile, 4 additional health care encounters per person-year versus no difference for men; at the 99^th^ percentile, 76 additional health care encounters per person-year for women, compared to 37 encounters per person-year for men), and women used more of all types of health care. For men, additional health care use occurred at the high end of utilization, where homecare use also decreased and there was no difference in emergency department use. A full understanding of why health care utilization differs by sex will be necessary to anticipate future long COVID health care resources.

Results of the sensitivity analysis requiring hospital discharge to start follow-up differed only by an expected decrease in the magnitude of additional days hospitalized per person-year among at the 99^th^ percentile of health care use. Although less than 1% of individuals entered long-term care, the relative impact of SARS-CoV-2 infection on mean days per person-year in long-term care was large, with a RR of 2.55 (95% CI 2.28-2.86; Table 3). While censoring follow-up at the date of entry to long-term care (Table E5) attenuated the magnitude of additional health care use for test-positive individuals, results were otherwise similar to main analyses with the exception of homecare among those at the 99^th^ percentile of utilization, which increased by 10.5 encounters per person-year. Due to barriers to entering to long-term care, which were exacerbated by the pandemic,^31^ our findings may underestimate the true need for long-term care associated with long COVID. Finally, findings were robust to also matching on intensive care unit admission within two weeks after the index date, with smaller magnitude differences but similar patterns in composite and component health care use.

Previous work indicates that 10-40% of individuals report symptoms months after acute COVID-19, including those who were not hospitalized initially, although these estimates vary by population and definitions of cases and outcomes.^46,47^ A subset of PCR test-positive individuals in Denmark who were contacted found that 53% reported symptoms 6-12 months after infection,^48^ and 5% of respondents in a study conducted in South Korea reported they were receiving treatment for symptoms a median of 454 days after their COVID-19 diagnosis.^49^ Several large studies conducted in the United States Veterans Health Administration health care system, ∼90% of whom were men and more than 70% were White, found increased outpatient clinic visits after hospitalization for COVID-19 or a positive SARS-CoV-2 PCR test, as well as increased risk of myocardial infarction and stroke in the months after infection; these findings may not be generalizable to other populations.^50,51^

### Limitations

Use of PCR results may misclassify infected individuals,^52,53^ although its relatively low sensitivity would be expected to bias results towards the null. Similarly, because cycle-time values for PCR tests were not available and viral cultures were not feasible, test-positive individuals may have been recently infected but no longer infectious at the time of their testing. Follow-up beginning eight weeks after testing was chosen to balance these considerations.

Using health care encounters as a measure of post-acute COVID-19 health care needs likely underestimates the true burden of long COVID, particularly as health care decreased during initial phases of the pandemic among some populations and in Ontario, in particular.^30,54-56^ There is no generally accepted method for weighing severity of different types of health care encounters^57,58^; our composite measure gives hospitalization and long-term care more weight due to their severity, although findings were robust in sensitivity analyses.

Because inclusion was conditioned on PCR testing, results may not generalize to populations with significant barriers to testing. Indication for testing and occupation (e.g., health care worker) were not available, so we are unable to determine whether these factors may modify associations between SARS-CoV-2 infection and type of post-acute health care use. To address potential changes in testing indications and capacity over time, test date was included in the propensity score and used for hard matching. Matching included multiple socio-demographic and clinical factors, although occupation, body mass index, or symptoms and their duration were not available. However, during the study period, testing was widely available for both symptomatic and asymptomatic individuals at no cost, reducing the risk of selection bias.

Results may not generalize to other variants of SARS-CoV-2 or populations with high prevalence of vaccination or re-infection,^59,60^ although they may be more applicable as new variants evade current vaccine coverage, previous vaccine immunity wanes, and public health protections are removed.^61^

### Conclusions

Post-acute health care utilization among patients with a positive SARS-CoV-2 PCR test is significantly higher compared to matched test-negative individuals, with higher rates of outpatient encounters, days hospitalized, and days in long-term care. Women had higher rates of health care use than men, particularly homecare. Given the number of infections worldwide, this translates to a tremendous increase in use of health care resources. Stakeholders can use these findings to prepare for long COVID health care demand.

## Data Availability

All data produced in the present work are contained in the manuscript

## Acknowledgments

This study was supported by ICES, which is funded in part by an annual grant from the Ontario Ministry of Health and the Ministry of Long-Term Care (MOHLTC). Part of this material is based on data and/or information compiled and provided by the Canadian Institute for Health Information (CIHI). The authors acknowledge that the clinical registry data used in this publications is from participating hospitals through CorHealth Ontario, which serves as an advisory body to the MOHLTC, is funded by the MOHLTC, and is dedicated to improving the quality, efficiency, access and equity in the delivery of the continuum of adult cardiac, vascular and stroke services in Ontario, Canada. The authors thank IQVIA Solutions Canada Inc. for use of their Drug Information File. Parts of this report are based on Ontario Registrar General (ORG) information on deaths, the original source of which is ServiceOntario. This study was supported by the Ontario Health Data Platform (OHDP), and Province of Ontario initiative to support Ontario’s ongoing response to COVID-19 and its related impacts. Parts of this material are based on data and/or information compiled and provided by CIHI and Cancer Care Ontario (CCO). The analyses, results, conclusions, opinions, and statements reported are those of the authors and are independent of the data and funding sources. No endorsements by ICES, the Ontario MOHLTC, CIHI, OHDP or its partners, ORG or the Ministry of Government Services, CCO, or the Province of Ontario is intended or should be inferred.

Johns Hopkins ACG® System Version 10 was used.

Dr. McNaughton is supported by the Sunnybrook Research Institute, the Practice Plan of the Department of Emergency Services at Sunnybrook Health Sciences Centre, and the University of Toronto.

Dr. Austin is supported by a Mid-Career Investigator Award from the Heart and Stroke Foundation.

Dr. Atzema is supported by the Sunnybrook Research Institute, the Practice Plan of the Department of Emergency Services at Sunnybrook Health Sciences Centre, and by a Mid-Career Investigator Award from the Heart and Stroke Foundation.

## Notes

### Competing Interest Statement

The authors have declared no competing interest.

### Funding Statement

This study was supported by ICES which is funded by an annual grant from the Ontario Ministry of Health (MOH). Please see the full document for additional funding details. CDM is supported by the Sunnybrook Research Institute and the Practice Plan of the Department of Emergency Services at Sunnybrook Health Sciences Centre and the University of Toronto.
PCA is supported by a Mid-Career Investigator Award from the Heart and Stroke Foundation. CLA is supported by the Sunnybrook Research Institute and the Practice Plan of the Department of Emergency Services at Sunnybrook Health Sciences Centre and by a Mid-Career Investigator Award from the Heart and Stroke Foundation.

### Author Declarations

ICES is a prescribed entity under Ontario's Personal Health Information Protection Act (PHIPA). Section 45 of PHIPA authorizes ICES to collect personal health information, without consent, for the purpose of analysis or compiling statistical information with respect to the management of, evaluation or monitoring of, the allocation of resources to or planning for all or part of the health system. Projects that use data collected by ICES under section 45 of PHIPA, and use no other data, are exempt from ethics committee/IRB review. The use of the data in this project is authorized under section 45 and approved by ICES' Privacy and Legal Office.

## References

1. Soriano JB, Murthy S, Marshall JC, Relan P, Diaz JV. W. H. O. Clinical Case Definition Working Group on Post-COVID-19 Condition. A clinical case definition of post-COVID-19 condition by a Delphi consensus. Lancet Infect Dis. 2021.

2. WHO Coronavirus (COVID-19) Dashboard. https://covid19.who.int. Last accessed January 14, 2022.

3. Fuhrmann J, Barbarossa MV. The significance of case detection ratios for predictions on the outcome of an epidemic - a message from mathematical modelers. Arch Public Health. 2020;78:63.

4. Greenhalgh T, Knight M, A’Court C, Buxton M, Husain L. Management of post-acute covid-19 in primary care. BMJ. 2020;370:m3026.

5. Xiao AT, Tong YX, Zhang S. False negative of RT-PCR and prolonged nucleic acid conversion in COVID-19: Rather than recurrence. J Med Virol. 2020.

6. Wu Y, Guo C, Tang L, et al. Prolonged presence of SARS-CoV-2 viral RNA in faecal samples. Lancet Gastroenterol Hepatol. 2020;5(5):434–435.

7. Raj SR, Arnold AC, Barboi A, et al. Long-COVID postural tachycardia syndrome: an American Autonomic Society statement. Clin Auton Res. 2021;31(3):365–368.

8. Tabacof L, Tosto-Mancuso J, Wood J, et al. Post-acute COVID-19 Syndrome Negatively Impacts Physical Function, Cognitive Function, Health-Related Quality of Life, and Participation. Am J Phys Med Rehabil. 2022;101(1):48–52.

9. Choi B, Choudhary MC, Regan J, et al. Persistence and Evolution of SARS-CoV-2 in an Immunocompromised Host. N Engl J Med. 2020;383(23):2291–2293.

10. Dennis A, Wamil M, Alberts J, et al. Multiorgan impairment in low-risk individuals with post-COVID-19 syndrome: a prospective, community-based study. BMJ open. 2021;11(3):e048391.

11. Donnelly JP, Wang XQ, Iwashyna TJ, Prescott HC. Readmission and Death After Initial Hospital Discharge Among Patients With COVID-19 in a Large Multihospital System. JAMA. 2021;325(3):304–306.

12. Zhou F, Yu T, Du R, et al. Clinical course and risk factors for mortality of adult inpatients with COVID-19 in Wuhan, China: a retrospective cohort study. Lancet. 2020.

13. Tenforde MW, Kim SS, Lindsell CJ, et al. Symptom Duration and Risk Factors for Delayed Return to Usual Health Among Outpatients with COVID-19 in a Multistate Health Care Systems Network - United States, March-June 2020. MMWR Morb Mortal Wkly Rep. 2020;69(30):993–998.

14. Ramchand R, Harrell MC, Berglass N, Lauck M. Veterans and COVID-19: Projecting the economic, social, and mental health needs of America’s Veterans. Bob Woodruff Foundation. March 2020. Accessed May 26, 2020. Bob Woodruff Foundation 2020.

15. Schull MJ, Azimaee M, Marra M, et al. ICES: Data, Discovery, Better Health. Int J Popul Data Sci. 2020;4(2):1135.

16. Austin PC. An Introduction to Propensity Score Methods for Reducing the Effects of Confounding in Observational Studies. Multivariate Behav Res. 2011;46(3):399–424.

17. Austin PC. Optimal caliper widths for propensity-score matching when estimating differences in means and differences in proportions in observational studies. Pharm Stat. 2011;10(2):150–161.

18. Austin PC. Balance diagnostics for comparing the distribution of baseline covariates between treatment groups in propensity-score matched samples. Stat Med. 2009;28(25):3083–3107.

19. Fernandez-de-Las-Penas C, Palacios-Cena D, Gomez-Mayordomo V, Cuadrado ML, Florencio LL. Defining Post-COVID Symptoms (Post-Acute COVID, Long COVID, Persistent Post-COVID): An Integrative Classification. Int J Environ Res Public Health. 2021;18(5).

20. Akbarialiabad H, Taghrir MH, Abdollahi A, et al. Long COVID, a comprehensive systematic scoping review. Infection. 2021;49(6):1163–1186.

21. Nalbandian A, Sehgal K, Gupta A, et al. Post-acute COVID-19 syndrome. Nat Med. 2021;27(4):601–615.

22. Tharakan T, Khoo CC, Giwercman A, et al. Are sex disparities in COVID-19 a predictable outcome of failing men’s health provision? Nat Rev Urol. 2022;19(1):47–63.

23. Gebhard C, Regitz-Zagrosek V, Neuhauser HK, Morgan R, Klein SL. Impact of sex and gender on COVID-19 outcomes in Europe. Biol Sex Differ. 2020;11(1):29.

24. Austin PC. Type I error rates, coverage of confidence intervals, and variance estimation in propensity-score matched analyses. Int J Biostat. 2009;5(1):Article 13.

25. Austin PC, Tu JV, Daly PA, Alter DA. The use of quantile regression in health care research: a case study examining gender differences in the timeliness of thrombolytic therapy. Stat Med. 2005;24(5):791–816.

26. Austin PC, Small DS. The use of bootstrapping when using propensity-score matching without replacement: a simulation study. Stat Med. 2014;33(24):4306–4319.

27. Khan JR, Awan N, Islam MM, Muurlink O. Healthcare Capacity, Health Expenditure, and Civil Society as Predictors of COVID-19 Case Fatalities: A Global Analysis. Front Public Health. 2020;8:347.

28. Jüni P, da Costa BR, Maltsev A, Katz GM, Perkhun A, Yan S, Bodmer NS. Ontario dashboard. Science Briefs of the Ontario COVID-19 Science Advisory Table. 2021. https://doi.org/10.47326/ocsat.dashboard.2021.1.0. Last accessed March 4, 2022.

29. Iuliano AD, Brunkard JM, Boehmer TK, et al. Trends in Disease Severity and Health Care Utilization During the Early Omicron Variant Period Compared with Previous SARS-CoV-2 High Transmission Periods - United States, December 2020-January 2022. MMWR Morb Mortal Wkly Rep. 2022;71(4):146–152.

30. Financial Accountability Office of Ontario. Ministry of Health: 2021 Spending Plan Review. https://www.fao-on.org/en/Blog/Publications/2021-health-estimates. Last accessed March 4, 2022.

31. Canadian Institute for Health Information. The Impact of COVID-19 on Long-Term Care in Canada: Focus on the First 6 Months. Ottawa, ON: CIHI; 2021.

32. Barrett KA, Vande Vyvere C, Haque N, et al. Critical care capacity during the COVID-19 pandemic. Science Briefs of the Ontario COVID-19 Science Advisory Table. 2021;2(51). https://doi.org/10.47326/ocsat.2021.02.51.1.0.

33. Brophy JT, Keith MM, Hurley M, McArthur JE. Sacrificed: Ontario Healthcare Workers in the Time of COVID-19. New Solut. 2021;30(4):267–281.

34. Cantor J, Whaley C, Simon K, Nguyen T. US Health Care Workforce Changes During the First and Second Years of the COVID-19 Pandemic. JAMA Health Forum. 2022;3(2):e215217–e215217.

35. Wilensky GR. The COVID-19 Pandemic and the US Health Care Workforce. JAMA Health Forum. 2022;3(1):e220001–e220001.

36. Wang J, Vahid S, Eberg M, et al. Clearing the surgical backlog caused by COVID-19 in Ontario: a time series modelling study. CMAJ. 2020;192(44):E1347–E1356.

37. Salenger R, Etchill EW, Ad N, et al. The Surge After the Surge: Cardiac Surgery Post-COVID-19. Ann Thorac Surg. 2020;110(6):2020–2025.

38. Bienvenu LA, Noonan J, Wang X, Peter K. Higher mortality of COVID-19 in males: sex differences in immune response and cardiovascular comorbidities. Cardiovasc Res. 2020;116(14):2197–2206.

39. Takahashi T, Ellingson MK, Wong P, et al. Sex differences in immune responses that underlie COVID-19 disease outcomes. Nature. 2020;588(7837):315–320.

40. Förster C, Colombo MG, Wetzel AJ, Martus P, Joos S. Persisting Symptoms After COVID-19-Prevalence and Risk Factors in a Population-Based Cohort. Dtsch Arztebl Int. 2022(Forthcoming).

41. Kashif A, Chaudhry M, Fayyaz T, et al. Follow-up of COVID-19 recovered patients with mild disease. Sci Rep. 2021;11(1):13414.

42. Bliddal S, Banasik K, Pedersen OB, et al. Acute and persistent symptoms in non-hospitalized PCR-confirmed COVID-19 patients. Sci Rep. 2021;11(1):13153.

43. Augustin M, Schommers P, Stecher M, et al. Post-COVID syndrome in non-hospitalised patients with COVID-19: a longitudinal prospective cohort study. Lancet Reg Health Eur. 2021;6:100122.

44. Stavem K, Ghanima W, Olsen MK, Gilboe HM, Einvik G. Persistent symptoms 1.5-6 months after COVID-19 in non-hospitalised subjects: a population-based cohort study. Thorax. 2021;76(4):405–407.

45. Wynberg E, van Willigen HDG, Dijkstra M, et al. Evolution of COVID-19 symptoms during the first 12 months after illness onset. Clin Infect Dis. 2021.

46. Smith MP. Estimating total morbidity burden of COVID-19: relative importance of death and disability. J Clin Epidemiol. 2022;142:54–59.

47. Cervia C, Zurbuchen Y, Taeschler P, et al. Immunoglobulin signature predicts risk of post-acute COVID-19 syndrome. Nat Commun. 2022;13(1):446.

48. Vedel Sørensen AI, Spiliopoulos L, Bager P, et al. Post-acute symptoms, new onset diagnoses and health problems 6 to 12 months after SARS-CoV-2 infection: a nationwide questionnaire study in the adult Danish population. medRxiv. 2022.

49. Kim Y, Bitna H, Kim SW, et al. Post-acute COVID-19 syndrome in patients after 12 months from COVID-19 infection in Korea. BMC Infect Dis. 2022;22(1):93.

50. Xie Y, Xu E, Bowe B, Al-Aly Z. Long-term cardiovascular outcomes of COVID-19. Nat Med. 2022.

51. Al-Aly Z, Xie Y, Bowe B. High-dimensional characterization of post-acute sequalae of COVID-19. Nature. 2021.

52. Axell-House DB, Lavingia R, Rafferty M, Clark E, Amirian ES, Chiao EY. The estimation of diagnostic accuracy of tests for COVID-19: A scoping review. J Infect. 2020;81(5):681–697.

53. Sundaram ME, Calzavara A, Mishra S, et al. Individual and social determinants of SARS-CoV-2 testing and positivity in Ontario, Canada: a population-wide study. CMAJ. 2021;193(20):E723–E734.

54. Howarth A, Munro M, Theodorou A, Mills PR. Trends in health care utilisation during COVID-19: a longitudinal study from the UK. BMJ open. 2021;11(7):e048151.

55. Moynihan R, Sanders S, Michaleff ZA, et al. Impact of COVID-19 pandemic on utilisation of health care services: a systematic review. BMJ open. 2021;11(3):e045343.

56. Mehrotra A, Bhatia RS, Snoswell CL. Paying for Telemedicine After the Pandemic. JAMA. 2021;325(5):431–432.

57. Wodchis WP, Austin PC, Henry DA. A 3-year study of high-cost users of health care. CMAJ. 2016;188(3):182–188.

58. Guilcher SJ, Bronskill SE, Guan J, Wodchis WP. Who Are the High-Cost Users? A Method for Person-Centred Attribution of Health Care Spending. PLoS One. 2016;11(3):e0149179.

59. Antonelli M, Penfold RS, Merino J, et al. Risk factors and disease profile of post-vaccination SARS-CoV-2 infection in UK users of the COVID Symptom Study app: a prospective, community-based, nested, case-control study. Lancet Infect Dis. 2022;22(1):43–55.

60. Tsuchida T, Hirose M, Inoue Y, Kunishima H, Otsubo T, Matsuda T. Relationship between changes in symptoms and antibody titers after a single vaccination in patients with Long COVID. J Med Virol. 2022.

61. Liu L, Iketani S, Guo Y, et al. Striking antibody evasion manifested by the Omicron variant of SARS-CoV-2. Nature. 2022;602(7898):676–681.

