## Supplemental materials for "Post-acute health care burden after SARS-CoV-2 infection: A retrospective cohort study of long COVID among 530,892 adults"

**Figure E1.** Cohort Construction

**Figure E2:** Among individuals at the 95th percentile of health care utilization, additional health care encounters (95% CI) per person-year total and by type of health care encounter, comparing individuals with a positive SARS-CoV-2 PCR test to matched individuals with negative PCR: (A) overall (n=530,232), (B) women (n=271,346), and (C) men (n=258,886).

**Table E1.** ICES Databases

**Table E2.** Variables included in the propensity score for matching

**Table E3**. Unmatched cohort baseline characteristics

**Table E4**. Distribution of baseline healthcare use by counts and rates (per person-year) for the matched cohort (n= 530,232), overall and stratified by sex.

**Table E5:** Sensitivity analyses: (A) Follow-up begins after hospital discharge or 56 days, whichever occurred later, (B) follow-up censored on the date of entrance to long-term care, (C) follow-up censored at 6 months, and (D) matched by intensive care admission within two weeks after index date. All comparisons are for test-positive versus test-negative individuals. Distribution of follow-up time included for each analysis. Healthcare utilization rates reported per person-year. The difference in overall healthcare utilization rates between test-positive and negative individuals are reported for the mean, 95th percentile, and 99th percentile. Samples sizes as noted below.

Figure E1: Cohort construction

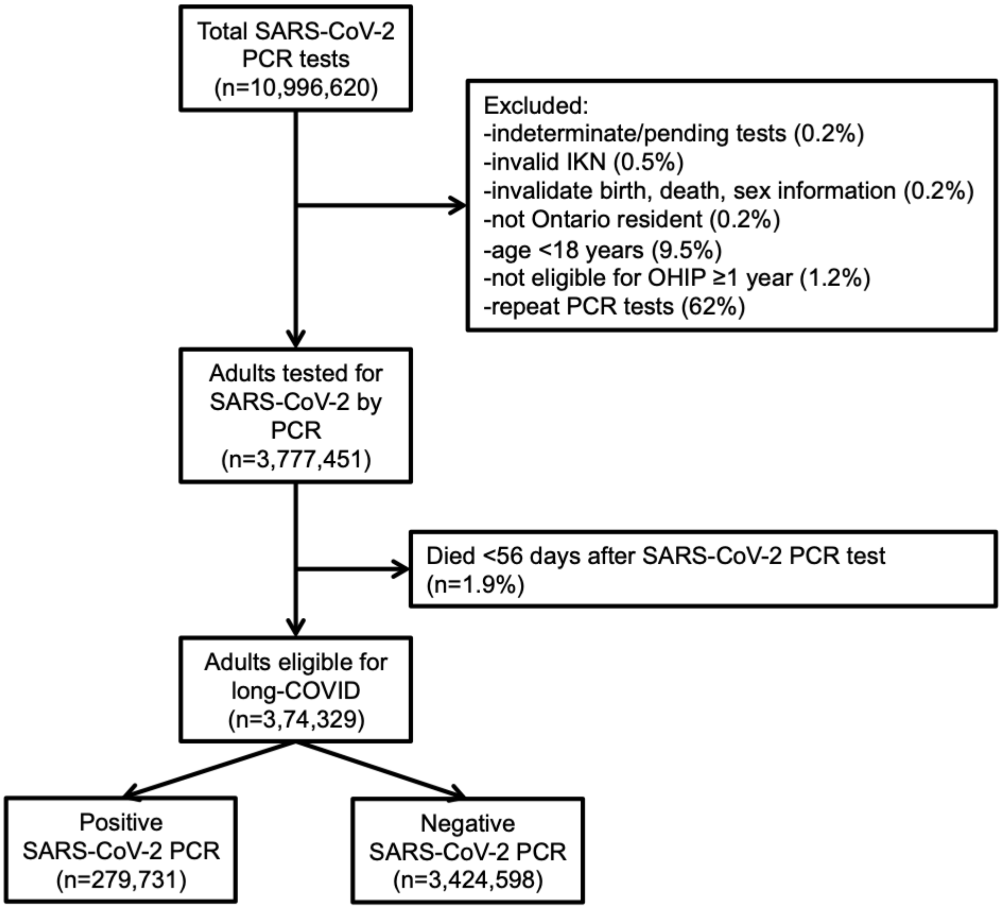

**Figure E2:** Among individuals at the 95th percentile of health care utilization, additional health care encounters (95% CI) per person-year total and by type of health care encounter, comparing individuals with a positive SARS-CoV-2 PCR test to matched individuals with negative PCR: (A) overall (n=530,232), (B) women (n=271,346), and (C) men (n=258,886).

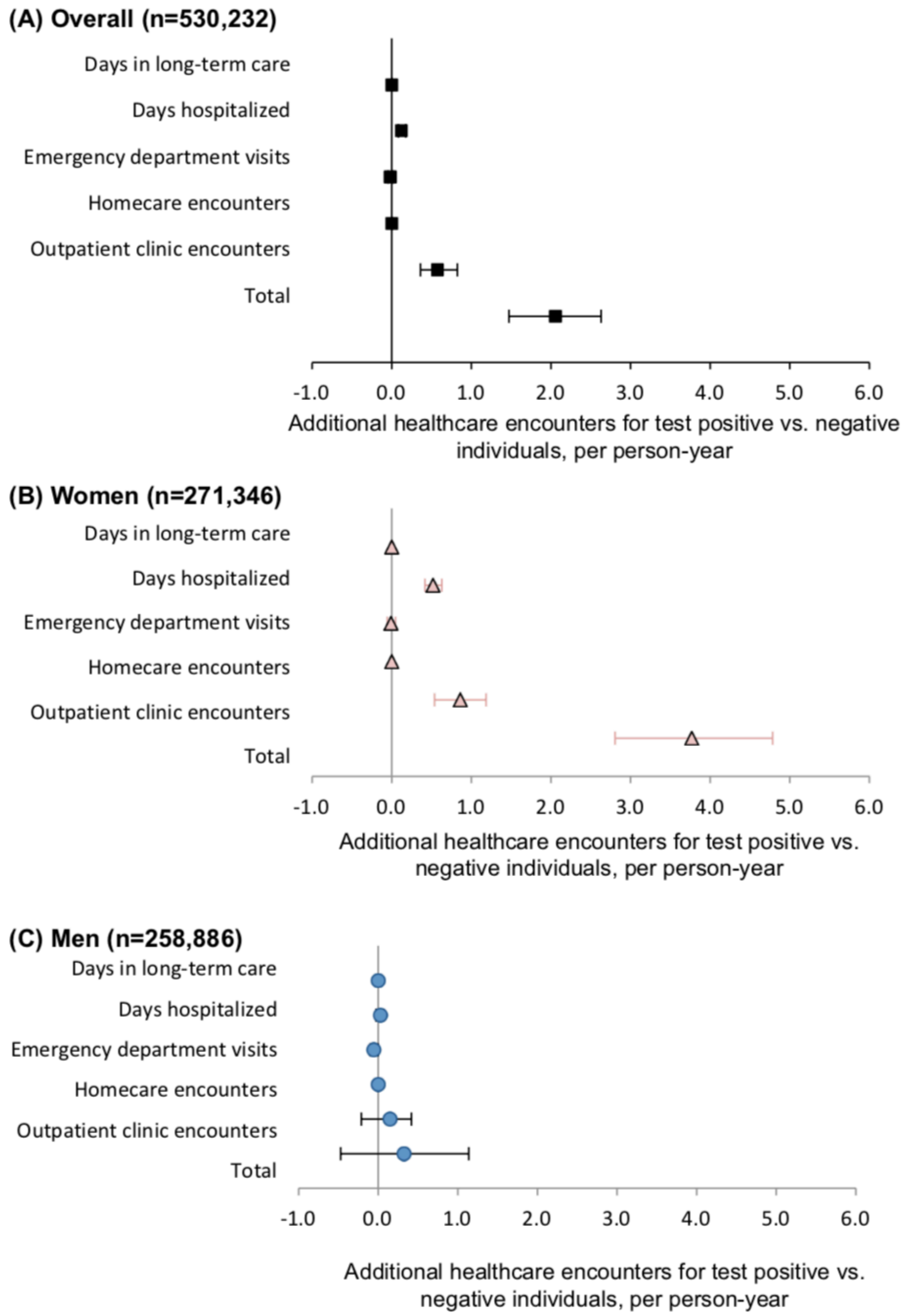

**Table E1.** ICES Databases
The following datasets were used for cohort construction, among others (Table E1): OHIP (contains all physician billing claims); the Registered Persons Database (maintains vital statistics, including out-of-hospital deaths); the Ontario Laboratories Information System (contains all PCR results for SARS-CoV-2 and was linked to the COVID19 database, which records all SARS-CoV-2 vaccinations); the Canadian Institute for Health Information Discharge Abstract Database (records all hospitalizations in Ontario); the National Ambulatory Care Reporting System (includes all emergency department visits in Ontario); and the Ministry of Health and Long-Term Care (MOHLTC; residence in long-term care).

| | | | | | |
|---|---|---|---|---|---|
| CIHI | NRS | HCD | CENSUS | INST | ODD |
| OHIP | ODB | RAICA | LIHN | ASTHMA | DEMENTIA |
| NACRS | CCRS | IPDB | REF | HYPER | COPD |
| SDS | CONTACT | CPDP | PCCF | HIV | ORAD |
| OHMRS | RAIHC | RPDB | DIN | CHF | OCCC |
| ODD | CCN | OCR | CDFP | ADP | ERCLAIM |
| AVGPRICE | ORGD | ETHNIC | Getacg | ESTSOB | GAPP |
| ONMARG | CCM | C19INTGR | DAD | COVAXON | OLIS |
| CAPE | NMS | OCCI | OHCAS | | |

**Table E2. Variables included in the propensity score for matching***
* also used for hard matching

| Variable | Definition and/or source |
|---|---|
| Sex* | Male, female |
| Week of outbreak* | Index date January 28, 2020; two-week blocks |
| Hospitalization* | On or within 2 weeks after the index date |
| Public health unit (PHU)* | OMHRS, DAD |
| Age | Years, restricted cubic spline |
| Neighbourhood income quintile | RPDB, PCCF |
| Residential instability | ONMARG quintiles |
| Material deprivation | ONMARG quintiles |
| Dependency | ONMARG quintiles |
| Ethnic concentration | ONMARG quintiles |
| Rurality | RPDB, PCCF |
| Week of outbreak | Index date January 28, 2020; two-week blocks |
| Diabetes | ODD_sensitive |
| Pregnancy | MOMBABY[1-4] |
| Hypertension | HYPER |
| Acute myocardial infarction | ≥1 DAD ICD10: I21, I22, I25.2 |
| Percutaneous coronary intervention | CIHI DAD/SDS: CCP: 48.02, 48.09; CCI: 1IJ50, 1IJ57GQ 1IJ80, 1IJ26, 1IJ54, 1IJ55; OHIP: Z434, Z448, Z449, Z460, Z461 |
| Coronary artery bypass surgery | (CIHI DAD/SDS): CCI 1IJ76; CCP 48.1, 48.2; OHIP R742, R743 |
| Ischemic stroke | One DAD, 2 OHIP, or 1 NACRS&1OHIP: ICD-10 codes I63, I64, H341 (excluding I63.6) as ANY diagnosis type, exclude suspect; ICD-9 code 434, 436 for OHIP |
| Hemorrhagic stroke | 1 record in DAD or NACRS: ICD-10 I60, I61; ICD-9 430, 431 |
| Major bleeding event | ICD 10:<br>GI: I850, I983, K250/252/254/256, K260/262/264/266, K270/272/274/276, K280/282/284/286, K290, K661, K920, K921, K922<br>ICH: I60, I61, I620, I621, I629<br>GU: N020-029, R310,R311, R318<br>Resp: R040, R041, R042, R048, R049<br>Other: R58, D68.3, H35.6, H45.0, M25.0 |

<table>
<tr><td>Solid cancer, hematologic cancer</td><td colspan="2">OCR: OCR_TOPOG_CD, OCR_DIAG_DATE</td></tr>
<tr><td>Same-day surgery in prior 6 weeks</td><td colspan="2">SDS_ADMDATE, SDS_INCODE1-10, SDS_CACSANETECH</td></tr>
<tr><td>Valvular disease</td><td colspan="2">ICD9: 394, 395, 396<br>ICD10: I019, I020, I05, I08, I099, I342, I348, I349<br>ICD10 code Z952, and CCI codes 1HS90LACF, 1HT90LACF, 1HU90DACF, 1HU90LACF, 1HU90PNCF, 1HV90LACF, 1HV90LACFA, 1HV90LACFL, 1HV90LACFN, 1HV90WJCFN</td></tr>
<tr><td>Emphysema</td><td colspan="2">COPD_SPECIFIC in COPD database</td></tr>
<tr><td>Asthma</td><td colspan="2">ASTHMA_SPEC in ASTHMA database</td></tr>
<tr><td>Atrial fibrillation</td><td colspan="2">Any of the following:<br>-history of hospitalization (CIHI DAD)[5]: ICD9 427.3 or ICD10 I48 as any diagnosis type, including suspected<br>-history of ED visit with same codes<br>-4 OHIP claims in 1 year (OHIP) dxcode 427</td></tr>
<tr><td>Heart failure</td><td colspan="2">CHF database</td></tr>
<tr><td>Ischemic heart disease</td><td colspan="2">PCI, CABG, (1 HOSP in DAD with any codes I20-I25) or (2 OHIP billings within a one-year period with dx codes 410-414)</td></tr>
<tr><td>Renal disease</td><td colspan="2">DAD code: ICD10 codes E102, E112, E132, E142, I12, I13, N01.*, N03.*, N05.*, N08.*, N18.*, N19.*, N25.*<br>or<br>Chronic Dialysis (Any 2 codes within 90 days of one another):<br>OHIP: R849, R850, G323, G325, G326, G330, G331, G860, G333, G083, G091, G085, G295, G082, G090, G092, G093, G094, G861, G862, G863, G864, G865, G866, G294, G095, G096 CCP: 51.95, 66.98<br>NACRS: CCI: 1PZ21HQBR, 1PZ21HPD4</td></tr>
<tr><td>Pneumonia</td><td colspan="2">(CIHI DAD, NACRS, OHIP) – ICD-10 codes J10.0, J11.0 or J12-J18 as ANY diagnosis type, exclude suspected[6]; OHIP dxcode 486, excluding claims associated with fee codes G538, G539, G840-G848, G590, G591 or G700 (administration of vaccinations)</td></tr>
<tr><td>Dementia</td><td colspan="2">(1 DAD or 3 OHIP billings separated by 30 days, within a 2-year period or any cholinesterase inhibitor from ODB)<br>ICD10 codes F00-F03, F051, G30, G31, R54<br>ICD9 code 290, 294, 331, 797<br>SUBCLNAM= CHOLINESTERASE INHIBITORS<br>ODB: donepezil, galantamine, or rivastigmine (DIN: 02232043, 02232044, 02269457, 02269465, 02244298, 02244299, 02244300, 02244302, 02266717, 02266725) or Tacrine (Cognex) (DIN: 66123288, 66123290, 66123306, 66123318)</td></tr>
<tr><td>Alcohol substance use disorder</td><td colspan="2">ICD10 codes F1094, F1029, F1019, F1099, F10250, F10150, F10950, F10920, F10929, F10251, F10251, F10151, F10951, F1027, F1097, F1026, F1096, F10129, F10120, F1010, F1021, F1020, F10220, F10229<br>ICD9 codes 291, 303</td></tr>
<tr><td>Johns Hopkins ACG score</td><td colspan="2">Sum of ADGs (from OHIP, NACRS-ED, CIHI DAD)</td></tr>
<tr><td>John Hopkins frailty indicator</td><td colspan="2">(OHIP, NACRS-ED CIHI DAD)</td></tr>
<tr><td>Influenza vaccination</td><td colspan="2">2019-20 influenza vaccination (OHIP, ODB) – Received between Sep 1, 2019 and COVID test date. Use algorithm in Concept Dictionary and attached MOHLTC bulletin (pdf) for publically funded DINs for 2019-20 season. Note: algorithm does not capture vaccinations received outside doctor's offices and pharmacies</td></tr>
<tr><td rowspan="8">Venous thromboembolism</td><td colspan="2">CIHI-NACRS, DAD, OHIP</td></tr>
<tr><td>ICD-10 Code</td><td>Description<br>DX10CODE1 in NACRS, DX10CODE1-10 in DAD</td></tr>
<tr><td>I26.*</td><td>Pulmonary embolism</td></tr>
<tr><td>I26.0</td><td>Pulmonary embolism with mention of acute cor pulmonale</td></tr>
<tr><td>I26.9</td><td>Pulmonary embolism without mention of acute cor pulmonale</td></tr>
<tr><td>I80.*</td><td>Phlebitis and thrombophlebitis[†]</td></tr>
<tr><td>I80.1</td><td>Phlebitis and thrombophlebitis of femoral vein</td></tr>
<tr><td>I80.2</td><td>Phlebitis and thrombophlebitis of other deep vessels of</td></tr>
</table>

| | | |
|---|---|---|
| | | lower extremities<br>- Deep vein thrombosis NOS |
| | I80.3 | Phlebitis and thrombophlebitis of lower extremities, unspecified<br>- Embolism or thrombosis of lower extremity NOS |
| | I80.8: | Phlebitis and thrombophlebitis of other sites; |
| | I80.9: | Phlebitis and thrombophlebitis of unspecified site |
| | I82.* | Other venous embolism and thrombosis‡ |
| | I82.2 | Embolism and thrombosis of vena cava |
| | I82.8 | Embolism and thrombosis of other specified veins |
| | I82.9 | Embolism and thrombosis of unspecified vein<br>-Embolism of vein NOS<br>-Thrombosis of vein NOS |
| | OHIP DXCODE | Description |
| | 415 | Pulmonary embolism, pulmonary infarction |
| | 451 | Phlebitis, thrombophlebitis |
| | 452 | Portal vein thrombosis |
| | 453 | Other venous embolism and thrombosis |
| | †Excludes: I80.0: Phlebitis and thrombophlebitis of superficial vessels of lower extremities<br>‡Excludes: I82.0: Budd-Chiari syndrome, I82.1: thrombophlebitis migrans. I82.3: Embolism and thrombosis of renal vein | |
| Mental health inpatient stay | NACRS | |
| Mental health ED visit | OHIP[7-9] | |
| MHA Outpatient Services | OHIP[7-9] | |
| Outpatient clinical encounters in previous year | OHIP | |
| ED visits in previous year | DAD, NACRS | |
| Same-day surgery in previous year | SDS_ADMDATE, SDS_INCODE1-10, SDS_CACSANETECH | |
| Days hospitalized in previous year | DAD | |
| Homecare encounters in previous year | HCD | |

**Supplemental Material References**


1. Aoyama K, Ray JG, Pinto R, et al. Temporal Variations in Incidence andOutcomes of Critical Illness Among Pregnant and Postpartum Women inCanada: A Population-Based Observational Study. *J Obstet Gynaecol Can.* 2019;41(5):631-640.
2. Metcalfe A, Lix LM, Johnson JA, et al. Validation of an obstetric comorbidity index in an external population. *BJOG.* 2015;122(13):1748-1755.
3. Joseph KS, Fahey J, Canadian Perinatal Surveillance S. Validation of perinatal data in the Discharge Abstract Database of the Canadian Institute for Health Information. *Chronic Dis Can.* 2009;29(3):96-100.
4. Samiedaluie S, Peterson S, Brant R, Kaczorowski J, Norman WV. Validating abortion procedure coding in Canadian administrative databases. *BMC Health Serv Res.* 2016;16:255.
5. Tu K, Nieuwlaat R, Cheng SY, et al. Identifying Patients With Atrial Fibrillation in Administrative Data. *Can J Cardiol.* 2016;32(12):1561-1565.
6. Griffin MR, Zhu Y, Moore MR, Whitney CG, Grijalva CG. U.S. hospitalizations for pneumonia after a decade of pneumococcal vaccination. *N Engl J Med.* 2013;369(2):155-163.
7. MHASEF Research Team. Mental Health and Addictions System Performance in Ontario: A Baseline Scorecard. Toronto, ON: Institute for Clinical Evaluative Sciences; 2018. Available from: https://www.ices.on.ca/Publications/Atlases-and-Reports/2018/MHASEF.
8. MHASEF Research Team. The Mental Health of Children and Youth in Ontario: 2017 Scorecard. Toronto, ON: Institute for Clinical Evaluative Sciences; 2017. ISBN: 978-1-926850-72-6. Available

from: https://www.ices.on.ca/Publications/Atlases-and-Reports/2017/MHASEF.

9. ICES Data Dictionary. https://datadictionary.ices.on.ca/Applications/DataDictionary/Default.aspx. Last accessed Feburary 11, 2022.

**Table E3:** Unmatched cohort, baseline characteristics

| VARIABLE | | Negative SARS-CoV-2 PCR N=3,362,519 | Positive SARS-CoV-2 PCR N=268,521 | Total N=3,631,040 | Standardized Difference |
|---|---|---|---|---|---|
| Age, years | Mean ± SD | 46.77 ± 18.17 | 44.18 ± 17.18 | 46.58 ± 18.11 | 0.15 |
| | Median (IQR) | 45 (31-60) | 43 (29-56) | 45 (31-60) | 0.14 |
| Women, n (%) | | 1,864,241 (55.4%) | 137,245 (51.1%) | 2,001,486 (55.1%) | 0.09 |
| Income Quintile, n (%) | Missing | 10,050 (0.3%) | 774 (0.3%) | 10,824 (0.3%) | 0 |
| | 1 | 630,532 (18.8%) | 66,584 (24.8%) | 697,116 (19.2%) | 0.15 |
| | 2 | 649,204 (19.3%) | 58,115 (21.6%) | 707,319 (19.5%) | 0.06 |
| | 3 | 672,162 (20.0%) | 57,233 (21.3%) | 729,395 (20.1%) | 0.03 |
| | 4 | 684,228 (20.3%) | 47,450 (17.7%) | 731,678 (20.2%) | 0.07 |
| | 5 | 716,343 (21.3%) | 38,365 (14.3%) | 754,708 (20.8%) | 0.18 |
| Instability Quintile, n (%) | Missing | 37,365 (1.1%) | 2,063 (0.8%) | 39,428 (1.1%) | 0.04 |
| | 1 | 691,223 (20.6%) | 72,327 (26.9%) | 763,550 (21.0%) | 0.15 |
| | 2 | 626,597 (18.6%) | 44,606 (16.6%) | 671,203 (18.5%) | 0.05 |
| | 3 | 610,179 (18.1%) | 40,798 (15.2%) | 650,977 (17.9%) | 0.08 |
| | 4 | 610,967 (18.2%) | 43,693 (16.3%) | 654,660 (18.0%) | 0.05 |
| | 5 | 786,188 (23.4%) | 65,034 (24.2%) | 851,222 (23.4%) | 0.02 |
| Deprivation Quintile, n (%) | Missing | 37,365 (1.1%) | 2,063 (0.8%) | 39,428 (1.1%) | 0.04 |
| | 1 | 802,901 (23.9%) | 45,106 (16.8%) | 848,007 (23.4%) | 0.18 |
| | 2 | 708,278 (21.1%) | 47,637 (17.7%) | 755,915 (20.8%) | 0.08 |
| | 3 | 632,209 (18.8%) | 52,504 (19.6%) | 684,713 (18.9%) | 0.02 |
| | 4 | 594,666 (17.7%) | 55,590 (20.7%) | 650,256 (17.9%) | 0.08 |
| | 5 | 587,100 (17.5%) | 65,621 (24.4%) | 652,721 (18.0%) | 0.17 |
| Dependency Quintile, n (%) | Missing | 37,365 (1.1%) | 2,063 (0.8%) | 39,428 (1.1%) | 0.04 |
| | 1 | 901,394 (26.8%) | 91,929 (34.2%) | 993,323 (27.4%) | 0.16 |
| | 2 | 671,373 (20.0%) | 59,930 (22.3%) | 731,303 (20.1%) | 0.06 |
| | 3 | 582,059 (17.3%) | 44,238 (16.5%) | 626,297 (17.2%) | 0.02 |
| | 4 | 557,097 (16.6%) | 37,506 (14.0%) | 594,603 (16.4%) | 0.07 |
| | 5 | 613,231 (18.2%) | 32,855 (12.2%) | 646,086 (17.8%) | 0.17 |
| Ethnic Concentration Quintile, n (%) | Missing | 37,365 (1.1%) | 2,063 (0.8%) | 39,428 (1.1%) | 0.04 |
| | 1 | 535,172 (15.9%) | 17,294 (6.4%) | 552,466 (15.2%) | 0.3 |
| | 2 | 579,980 (17.2%) | 24,901 (9.3%) | 604,881 (16.7%) | 0.24 |
| | 3 | 635,538 (18.9%) | 35,134 (13.1%) | 670,672 (18.5%) | 0.16 |
| | 4 | 726,900 (21.6%) | 57,068 (21.3%) | 783,968 (21.6%) | 0.01 |
| | 5 | 847,564 (25.2%) | 132,061 (49.2%) | 979,625 (27.0%) | 0.51 |
| Rural, n (%) | Missing | 8,780 (0.3%) | 671 (0.2%) | 9,451 (0.3%) | 0 |
| | | 339,985 (10.1%) | 10,538 (3.9%) | 350,523 (9.7%) | 0.24 |
| Pandemic Quarter | 2020-Q1 | 19,479 (0.6%) | 3,457 (1.3%) | 22,936 (0.6%) | 0.07 |
| | 2020-Q2 | 297,938 (8.9%) | 21,179 (7.9%) | 319,117 (8.8%) | 0.04 |
| | 2020-Q3 | 686,688 (20.4%) | 14,775 (5.5%) | 701,463 (19.3%) | 0.46 |
| | 2020-Q4 | 972,169 (28.9%) | 104,177 (38.8%) | 1,076,346 (29.6%) | 0.21 |
| | 2021-Q1 | 1,386,245 (41.2%) | 124,933 (46.5%) | 1,511,178 (41.6%) | 0.11 |
| Received 2 vaccine doses, n (%) | | 71,870 (2.1%) | 300 (0.1%) | 72,170 (2.0%) | 0.19 |
| Received 1 vaccine dose, n (%) | | 78,444 (2.3%) | 1,403 (0.5%) | 79,847 (2.2%) | 0.15 |
| Received 0 vaccine doses, n (%) | | 3,212,205 (95.5%) | 266,818 (99.4%) | 3,479,023 (95.8%) | 0.25 |

| | | | | | |
|---|---|---|---|---|---|
| Aggregated diagnosis group | Mean ± SD | 5.82 ± 3.80 | 5.59 ± 3.69 | 5.81 ± 3.79 | 0.06 |
| | Median (IQR) | 5 (3-8) | 5 (3-8) | 5 (3-8) | 0.06 |
| Hospital Frailty Risk Score | Mean ± SD | 2.42 ± 4.76 | 2.40 ± 5.03 | 2.42 ± 4.78 | 0 |
| | Median (IQR) | 0 (0-3) | 0 (0-2) | 0 (0-3) | 0.06 |
| Hospitalizations in prior year | Mean ± SD | 0.10 ± 0.45 | 0.07 ± 0.39 | 0.10 ± 0.44 | 0.07 |
| | Median (IQR) | 0 (0-0) | 0 (0-0) | 0 (0-0) | 0.09 |
| Clinic visits in prior year | Mean ± SD | 6.56 ± 8.35 | 6.27 ± 7.86 | 6.54 ± 8.32 | 0.04 |
| | Median (IQR) | 4 (1-9) | 4 (1-9) | 4 (1-9) | 0.03 |
| Homecare visits in prior year | Mean ± SD | 3.10 ± 25.71 | 2.92 ± 26.23 | 3.09 ± 25.75 | 0.01 |
| | Median (IQR) | 0 (0-0) | 0 (0-0) | 0 (0-0) | 0.07 |
| ED visits in prior year | Mean ± SD | 0.50 ± 1.45 | 0.41 ± 1.42 | 0.49 ± 1.45 | 0.06 |
| | Median (IQR) | 0 (0-1) | 0 (0-0) | 0 (0-1) | 0.08 |
| Days hospitalized in prior year | Mean ± SD | 1.02 ± 6.85 | 0.83 ± 7.88 | 1.00 ± 6.93 | 0.03 |
| | Median (IQR) | 0 (0-0) | 0 (0-0) | 0 (0-0) | 0.15 |
| Hospitalized within 2 weeks, n (%) | | 246,238 (7.3%) | 14,942 (5.6%) | 261,180 (7.2%) | 0.07 |
| Admitted to intensive care unit within 2 weeks, n (%) | | 31,068 (0.9%) | 2,933 (1.1%) | 34,001 (0.9%) | 0.02 |
| Johns Hopkins Frailty Index, n (%) | | 116,104 (3.5%) | 7,895 (2.9%) | 123,999 (3.4%) | 0.03 |
| Flu vaccine within prior year, n (%) | | 1,064,518 (31.7%) | 66,351 (24.7%) | 1,130,869 (31.1%) | 0.15 |
| Pregnancy, n (%) | | 29,558 (0.9%) | 1,855 (0.7%) | 31,413 (0.9%) | 0.02 |
| Hypertension, n (%) | | 816,144 (24.3%) | 61,708 (23.0%) | 877,852 (24.2%) | 0.03 |
| Diabetes, n (%) | | 395,575 (11.8%) | 37,775 (14.1%) | 433,350 (11.9%) | 0.07 |
| Emphysema, n (%) | | 93,686 (2.8%) | 4,138 (1.5%) | 97,824 (2.7%) | 0.09 |
| Heart failure, n (%) | | 88,669 (2.6%) | 4,994 (1.9%) | 93,663 (2.6%) | 0.05 |
| Dementia, n (%) | | 42,410 (1.3%) | 3,274 (1.2%) | 45,684 (1.3%) | 0 |
| Asthma, n (%) | | 414,612 (12.3%) | 28,270 (10.5%) | 442,882 (12.2%) | 0.06 |
| Cancer, n (%) | | 112,188 (3.3%) | 4,809 (1.8%) | 116,997 (3.2%) | 0.1 |
| Surgery in prior 6 weeks, n (%) | | 57,709 (1.7%) | 2,215 (0.8%) | 59,924 (1.7%) | 0.08 |
| Ischemic stroke, n (%) | | 42,908 (1.3%) | 2,547 (0.9%) | 45,455 (1.3%) | 0.03 |
| Hemorrhagic stroke, n (%) | | 3,533 (0.1%) | 224 (0.1%) | 3,757 (0.1%) | 0.01 |
| Valvular disease, n (%) | | 4,609 (0.1%) | 235 (0.1%) | 4,844 (0.1%) | 0.01 |
| Atrial fibrillation, n (%) | | 93,557 (2.8%) | 4,896 (1.8%) | 98,453 (2.7%) | 0.06 |
| Myocardial infarction, n (%) | | 33,446 (1.0%) | 1,801 (0.7%) | 35,247 (1.0%) | 0.04 |
| Percutaneous coronary intervention, n (%) | | 39,373 (1.2%) | 2,231 (0.8%) | 41,604 (1.1%) | 0.03 |
| Coronary artery bypass, n (%) | | 11,845 (0.4%) | 648 (0.2%) | 12,493 (0.3%) | 0.02 |
| Ischemic heart disease, n (%) | | 167,514 (5.0%) | 9,846 (3.7%) | 177,360 (4.9%) | 0.06 |
| Major bleeding, n (%) | | 37,210 (1.1%) | 2,156 (0.8%) | 39,366 (1.1%) | 0.03 |
| Renal disease, n (%) | | 43,949 (1.3%) | 2,779 (1.0%) | 46,728 (1.3%) | 0.03 |
| Pneumonia, n (%) | | 271,036 (8.1%) | 20,166 (7.5%) | 291,202 (8.0%) | 0.02 |
| Alcohol use disorder, n (%) | | 27,759 (0.8%) | 1,668 (0.6%) | 29,427 (0.8%) | 0.02 |
| Venous thromboembolism, n (%) | | 396,371 (11.8%) | 24,085 (9.0%) | 420,456 (11.6%) | 0.09 |
| Mental health hospitalization, n (%) | | 77,945 (2.3%) | 4,470 (1.7%) | 82,415 (2.3%) | 0.05 |
| Mental health emergency visit, n (%) | | 188,832 (5.6%) | 11,469 (4.3%) | 200,301 (5.5%) | 0.06 |
| Mental health clinic visit, n (%) | | 747,646 (22.2%) | 48,712 (18.1%) | 796,358 (21.9%) | 0.1 |
| Hospital mental health diagnoses, no. (%) | | | | | |

| | | | | |
|---|---|---|---|---|
| Substance use disorder | 19,681 (0.6%) | 1,138 (0.4%) | 20,819 (0.6%) | 0.02 |
| Dementia/Alzheimer's | 2,034 (0.1%) | 193 (0.1%) | 2,227 (0.1%) | 0 |
| Delirium | 5,696 (0.2%) | 423 (0.2%) | 6,119 (0.2%) | 0 |
| Anxiety | 4,340 (0.1%) | 203 (0.1%) | 4,543 (0.1%) | 0.02 |
| Deliberate self-harm | 5,492 (0.2%) | 354 (0.1%) | 5,846 (0.2%) | 0.01 |
| Schizophrenia | 14,352 (0.4%) | 937 (0.3%) | 15,289 (0.4%) | 0.01 |
| Mood disorders | 636 (0.0%) | 23 (0.0%) | 659 (0.0%) | 0.01 |
| Mood disorder - bipolar | 8,380 (0.2%) | 417 (0.2%) | 8,797 (0.2%) | 0.02 |
| Mood disorder - depression | 21,114 (0.6%) | 1,015 (0.4%) | 22,129 (0.6%) | 0.04 |
| Mood disorder - other | 347 (0.0%) | 12 (0.0%) | 359 (0.0%) | 0.01 |
| Personality disorders | 5,683 (0.2%) | 234 (0.1%) | 5,917 (0.2%) | 0.02 |
| Nonclassified | 4,919 (0.1%) | 302 (0.1%) | 5,221 (0.1%) | 0.01 |
| Trauma/stress related disorders | 9,008 (0.3%) | 443 (0.2%) | 9,451 (0.3%) | 0.02 |
| Obsessive compulsive disorder | 465 (0.0%) | 26 (0.0%) | 491 (0.0%) | 0 |
| Eating disorder | 854 (0.0%) | 37 (0.0%) | 891 (0.0%) | 0.01 |
| Emergency department mental health diagnoses, no. (%) | | | | |
| Substance use disorder | 60,371 (1.8%) | 4,178 (1.6%) | 64,549 (1.8%) | 0.02 |
| Dementia/Alzheimer's | 1,971 (0.1%) | 158 (0.1%) | 2,129 (0.1%) | 0 |
| Delirium | 1,195 (0.0%) | 72 (0.0%) | 1,267 (0.0%) | 0 |
| Anxiety | 68,125 (2.0%) | 3,872 (1.4%) | 71,997 (2.0%) | 0.04 |
| Deliberate self-harm | 16,222 (0.5%) | 1,057 (0.4%) | 17,279 (0.5%) | 0.01 |
| Schizophrenia | 11,013 (0.3%) | 745 (0.3%) | 11,758 (0.3%) | 0.01 |
| Bipolar | 5,640 (0.2%) | 262 (0.1%) | 5,902 (0.2%) | 0.02 |
| Depression | 34,030 (1.0%) | 1,778 (0.7%) | 35,808 (1.0%) | 0.04 |
| Other | 2,265 (0.1%) | 108 (0.0%) | 2,373 (0.1%) | 0.01 |
| Personality disorders | 8,318 (0.2%) | 446 (0.2%) | 8,764 (0.2%) | 0.02 |
| Nonclassified | 6,950 (0.2%) | 473 (0.2%) | 7,423 (0.2%) | 0.01 |
| Trauma/stress related disorders | 41,958 (1.2%) | 2,202 (0.8%) | 44,160 (1.2%) | 0.04 |
| Obsessive compulsive disorder | 626 (0.0%) | 39 (0.0%) | 665 (0.0%) | 0 |
| Eating disorder | 614 (0.0%) | 22 (0.0%) | 636 (0.0%) | 0.01 |
| Outpatient OHIP mental health diagnosis codes, no. (%) | | | | |
| Psychotic disorders | 43,698 (1.3%) | 3,137 (1.2%) | 46,835 (1.3%) | 0.01 |
| Mood and anxiety disorders | 385,325 (11.5%) | 22,799 (8.5%) | 408,124 (11.2%) | 0.1 |
| Substance use disorders | 110,162 (3.3%) | 6,198 (2.3%) | 116,360 (3.2%) | 0.06 |
| Non-psychotic disorders | 190,835 (5.7%) | 12,795 (4.8%) | 203,630 (5.6%) | 0.04 |
| Social problems | 127,299 (3.8%) | 8,967 (3.3%) | 136,266 (3.8%) | 0.02 |
| Other | 139,737 (4.2%) | 9,465 (3.5%) | 149,202 (4.1%) | 0.03 |

**Table E4:** Distribution of baseline healthcare utilization rates (per person-year) for the matched cohort (n= 530,232), overall and stratified by sex.

| Rate of baseline healthcare utilization (previous year; per person year) | SARS-CoV-2 PCR Test Result | Mean | std | Q1 | Median | Q3 | p95 | p99 |
|---|---|---|---|---|---|---|---|---|
| **Overall** | | | | | | | | |
| Outpatient clinical encounters | Negative | 6.3 | 7.9 | 1.0 | 4.0 | 8.0 | 21.0 | 37.0 |
| | Positive | 6.3 | 7.8 | 1.0 | 4.0 | 8.0 | 21.0 | 36.0 |
| Homecare encounters | Negative | 2.7 | 25.0 | 0.0 | 0.0 | 0.0 | 0.0 | 91.1 |
| | Positive | 2.8 | 25.7 | 0.0 | 0.0 | 0.0 | 0.0 | 95.1 |
| Emergency department visits | Negative | 0.4 | 1.2 | 0.0 | 0.0 | 0.0 | 2.0 | 4.0 |
| | Positive | 0.4 | 1.4 | 0.0 | 0.0 | 0.0 | 2.0 | 4.0 |
| Days hospitalized | Negative | 0.8 | 6.9 | 0.0 | 0.0 | 0.0 | 2.0 | 17.0 |
| | Positive | 0.8 | 7.3 | 0.0 | 0.0 | 0.0 | 2.0 | 17.0 |
| **Total healthcare utilization** | Negative | 10.2 | 29.5 | 1.0 | 4.0 | 10.0 | 29.0 | 131.1 |
| | Positive | 10.3 | 30.3 | 1.0 | 4.0 | 10.0 | 28.0 | 137.1 |
| **Women (n=271,346)** | | | | | | | | |
| Outpatient clinical encounters | Negative | 7.2 | 8.3 | 2.0 | 5.0 | 10.0 | 23.0 | 38.0 |
| | Positive | 7.3 | 8.2 | 2.0 | 5.0 | 10.0 | 23.0 | 38.0 |
| Homecare encounters | Negative | 3.1 | 26.8 | 0.0 | 0.0 | 0.0 | 0.0 | 104.1 |
| | Positive | 3.5 | 29.4 | 0.0 | 0.0 | 0.0 | 0.0 | 132.1 |
| Emergency department visits | Negative | 0.4 | 1.1 | 0.0 | 0.0 | 0.0 | 2.0 | 4.0 |
| | Positive | 0.4 | 1.2 | 0.0 | 0.0 | 0.0 | 2.0 | 4.0 |
| Days hospitalized | Negative | 0.7 | 6.2 | 0.0 | 0.0 | 0.0 | 2.0 | 15.0 |
| | Positive | 0.8 | 6.4 | 0.0 | 0.0 | 0.0 | 2.0 | 16.0 |
| **Men (n=258,886)** | | | | | | | | |
| Outpatient clinical encounters | Negative | 5.3 | 7.4 | 1.0 | 3.0 | 7.0 | 19.0 | 35.0 |
| | Positive | 5.2 | 7.4 | 1.0 | 3.0 | 7.0 | 18.0 | 34.0 |
| Homecare encounters | Negative | 2.4 | 22.8 | 0.0 | 0.0 | 0.0 | 0.0 | 68.0 |
| | Positive | 2.0 | 21.1 | 0.0 | 0.0 | 0.0 | 0.0 | 50.0 |
| Emergency department visits | Negative | 0.4 | 1.2 | 0.0 | 0.0 | 0.0 | 2.0 | 4.0 |
| | Positive | 0.4 | 1.5 | 0.0 | 0.0 | 0.0 | 2.0 | 4.0 |
| Days hospitalized | Negative | 0.9 | 7.6 | 0.0 | 0.0 | 0.0 | 2.0 | 20.0 |
| | Positive | 0.8 | 8.1 | 0.0 | 0.0 | 0.0 | 1.0 | 18.0 |

**Table E5:** Sensitivity analyses: (A) Follow-up begins after hospital discharge or 56 days, whichever occurred later, (B) follow-up censored on the date of entrance to long-term care, (C) follow-up censored at 6 months, and (D) matched by intensive care admission within two weeks after index date. All comparisons are for test-positive versus test-negative individuals. Distribution of follow-up time included for each analysis. Healthcare utilization rates reported per person-year. The difference in overall healthcare utilization rates between test-positive and negative individuals are reported for the mean, 95th percentile, and 99th percentile. Samples sizes as noted below.

| Rates (per person-year) | SARS-CoV-2 Positive PCR Test Result | Mean | std | ∆ for mean | Q1 | Median | Q3 | p95 | p99 | ∆ for 95th percentile | ∆ for 99th percentile |
|---|---|---|---|---|---|---|---|---|---|---|---|
| **Sensitivity analysis (A): Follow-up ≥56 days (after hospital discharge) (n=530,232)** | | | | | | | | | | | |
| Total healthcare utilization | Negative | 12.8 | 40.5 | | 1.2 | 4.5 | 10.7 | 34.5 | 264.4 | | |
| | Positive | 14.2 | 45.1 | 1.3 | 1.4 | 4.9 | 11.3 | 36.4 | 335.4 | 1.8 | 71.0 |
| Outpatient clinical encounters | Negative | 7.1 | 10.6 | | 0.0 | 4.0 | 9.5 | 24.7 | 45.1 | | |
| | Positive | 7.4 | 11.2 | 0.3 | 1.0 | 4.4 | 10.1 | 25.2 | 45.0 | 0.6 | -0.1 |
| Homecare encounters | Negative | 4.1 | 32.4 | | 0.0 | 0.0 | 0.0 | 0.0 | 167.4 | | |
| | Positive | 4.2 | 32.5 | 0.0 | 0.0 | 0.0 | 0.0 | 0.0 | 169.6 | 0.0 | 2.2 |
| Emergency department visits | Negative | 0.4 | 1.5 | | 0.0 | 0.0 | 0.0 | 2.5 | 5.4 | | |
| | Positive | 0.4 | 1.7 | 0.0 | 0.0 | 0.0 | 0.0 | 2.4 | 5.5 | 0.0 | 0.1 |
| Days hospitalized | Negative | 0.7 | 8.3 | | 0.0 | 0.0 | 0.0 | 1.7 | 13.8 | | |
| | Positive | 1.0 | 11.7 | 0.3 | 0.0 | 0.0 | 0.0 | 1.8 | 16.0 | 0.1 | 2.2 |
| Days in long-term care | Negative | 0.5 | 11.6 | | 0.0 | 0.0 | 0.0 | 0.0 | 0.0 | | |
| | Positive | 1.2 | 19.2 | 0.7 | 0.0 | 0.0 | 0.0 | 0.0 | 0.0 | 0.0 | 0.0 |
| **Follow-up (days)** | Negative | 240 | 87 | | 187 | 221 | 267 | 457 | 495 | | |
| | Positive | 240 | 87 | | 188 | 221 | 267 | 457 | 494 | | |
| **Sensitivity analysis (B): Follow-up censored at entrance to long-term care (n=530,232)** | | | | | | | | | | | |
| **Rates (per person-year)** | | **Mean** | **std** | **∆ for mean** | **Q1** | **Median** | **Q3** | **p95** | **p99** | **∆ for 95th percentile** | **∆ for 99th percentile** |
| Total healthcare utilization | Negative | 12.5 | 39.2 | | 1.2 | 4.5 | 10.7 | 34.2 | 243.5 | | |
| | Positive | 13.6 | 42.5 | 1.1 | 1.4 | 4.9 | 11.2 | 35.7 | 297.5 | 1.5 | 54.0 |
| Outpatient clinical encounters | Negative | 7.1 | 10.4 | | 0.0 | 4.0 | 9.6 | 24.7 | 45.2 | | |
| | Positive | 7.4 | 10.7 | 0.3 | 1.0 | 4.4 | 10.1 | 25.4 | 45.2 | 0.7 | 0.0 |
| Homecare encounters | Negative | 4.2 | 32.8 | | 0.0 | 0.0 | 0.0 | 0.0 | 172.1 | | |
| | Positive | 4.3 | 33.3 | 0.1 | 0.0 | 0.0 | 0.0 | 0.0 | 182.6 | 0.0 | 10.5 |
| Emergency department visits | Negative | 0.4 | 1.5 | | 0.0 | 0.0 | 0.0 | 2.5 | 5.4 | | |
| | Positive | 0.4 | 1.7 | 0.0 | 0.0 | 0.0 | 0.0 | 2.4 | 5.5 | 0.0 | 0.1 |
| Days hospitalized | Negative | 0.8 | 10.1 | | 0.0 | 0.0 | 0.0 | 1.7 | 14.5 | | |
| | Positive | 1.5 | 16.7 | 0.6 | 0.0 | 0.0 | 0.0 | 1.9 | 22.1 | 0.1 | 7.6 |
| Days in long-term care | Negative | . | . | | . | . | . | . | . | | |
| | Positive | . | . | | . | . | . | . | . | | |
| **Follow-up (days)** | Negative | 240 | 87 | | 186 | 221 | 267 | 457 | 495 | | |
| | Positive | 239 | 88 | | 187 | 220 | 267 | 457 | 494 | | |
| **Sensitivity analysis (C) Follow-up censored at 6 months (n=530,232)** | | | | | | | | | | | |
| **Rates (per person-year)** | | **Mean** | **std** | **∆ for mean** | **Q1** | **Median** | **Q3** | **p95** | **p99** | **∆ for 95th percentile** | **∆ for 99th percentile** |
| Total healthcare utilization | Negative | 12.8 | 40.6 | | 0.0 | 4.0 | 10.1 | 35.1 | 264.4 | | |
| | Positive | 14.3 | 45.0 | 4.4 | 0.0 | 4.0 | 12.1 | 38.3 | 337.0 | 3.2 | 72.6 |
| Outpatient clinical encounters | Negative | 7.1 | 10.7 | | 0.0 | 4.0 | 10.1 | 26.2 | 46.4 | | |

| | | | | | | | | | | | |
|---|---|---|---|---|---|---|---|---|---|---|---|
| | Positive | 7.5 | 11.0 | 0.3 | 0.0 | 4.0 | 10.1 | 26.2 | 46.4 | 0.0 | 0.0 |
| Homecare encounters | Negative | 4.1 | 32.6 | | 0.0 | 0.0 | 0.0 | 0.0 | 167.5 | | |
| | Positive | 4.1 | 32.5 | -0.1 | 0.0 | 0.0 | 0.0 | 0.0 | 163.5 | 0.0 | -4.0 |
| Emergency department visits | Negative | 0.4 | 1.5 | | 0.0 | 0.0 | 0.0 | 2.2 | 6.1 | | |
| | Positive | 0.4 | 1.7 | 0.2 | 0.0 | 0.0 | 0.0 | 2.1 | 6.1 | 0.0 | 0.0 |
| Days hospitalized | Negative | 0.8 | 9.0 | | 0.0 | 0.0 | 0.0 | 2.0 | 14.1 | | |
| | Positive | 1.3 | 13.7 | 4.7 | 0.0 | 0.0 | 0.0 | 2.0 | 22.2 | 0.0 | 8.1 |
| Days in long-term care | Negative | 0.4 | 10.7 | | 0.0 | 0.0 | 0.0 | 0.0 | 0.0 | | |
| | Positive | 1.0 | 17.2 | 6.4 | 0.0 | 0.0 | 0.0 | 0.0 | 0.0 | 0.0 | 0.0 |
| **Follow-up (days)** | Negative | 174 | 17 | | 181 | 181 | 181 | 181 | 181 | | |
| | Positive | 174 | 17 | | 181 | 181 | 181 | 181 | 181 | | |

**Sensitivity analysis (D) Matched by intensive care unit admission within 2 weeks after index date (n=508,662)**

| **Rates (per person-year)** | | **Mean** | **std** | **∆ for mean** | **Q1** | **Median** | **Q3** | **p95** | **p99** | **∆ for 95th percentile** | **∆ for 99th percentile** |
|---|---|---|---|---|---|---|---|---|---|---|---|
| Total healthcare utilization | Negative | 12.8 | 40.6 | | 1.1 | 4.5 | 10.7 | 34.5 | 268.1 | | |
| | Positive | 14.1 | 44.6 | 1.3 | 1.4 | 4.9 | 11.2 | 36.2 | 332.5 | 1.7 | 64.4 |
| Outpatient clinical encounters | Negative | 7.1 | 10.3 | | 0.0 | 4.0 | 9.5 | 24.6 | 45.3 | | |
| | Positive | 7.4 | 10.6 | 0.3 | 1.0 | 4.4 | 10.0 | 25.2 | 44.9 | 0.6 | -0.4 |
| Homecare encounters | Negative | 4.1 | 32.4 | | 0.0 | 0.0 | 0.0 | 0.0 | 163.5 | | |
| | Positive | 4.1 | 32.0 | 0.0 | 0.0 | 0.0 | 0.0 | 0.0 | 161.4 | 0.0 | -2.2 |
| Emergency department visits | Negative | 0.4 | 1.5 | | 0.0 | 0.0 | 0.0 | 2.5 | 5.4 | | |
| | Positive | 0.4 | 1.7 | 0.0 | 0.0 | 0.0 | 0.0 | 2.4 | 5.5 | 0.0 | 0.0 |
| Days hospitalized | Negative | 0.8 | 8.8 | | 0.0 | 0.0 | 0.0 | 1.7 | 13.9 | | |
| | Positive | 1.1 | 12.4 | 0.4 | 0.0 | 0.0 | 0.0 | 1.8 | 20.1 | 0.1 | 6.2 |
| Days in long-term care | Negative | 0.5 | 11.7 | | 0.0 | 0.0 | 0.0 | 0.0 | 0.0 | | |
| | Positive | 1.1 | 18.2 | 0.6 | 0.0 | 0.0 | 0.0 | 0.0 | 0.0 | 0.0 | 0.0 |
